# Measurable imaging-based changes in enhancement of intrahepatic cholangiocarcinoma after radiotherapy reflect physical mechanisms of response

**DOI:** 10.1101/2024.09.11.24313334

**Authors:** Brian De, Prashant Dogra, Mohamed Zaid, Dalia Elganainy, Kevin Sun, Ahmed M. Amer, Charles Wang, Michael K. Rooney, Enoch Chang, Hyunseon C. Kang, Zhihui Wang, Priya Bhosale, Bruno C. Odisio, Timothy E. Newhook, Ching-Wei D. Tzeng, Hop S. Tran Cao, Yun S. Chun, Jean-Nicholas Vauthey, Sunyoung S. Lee, Ahmed Kaseb, Kanwal Raghav, Milind Javle, Bruce D. Minsky, Sonal S. Noticewala, Emma B. Holliday, Grace L. Smith, Albert C. Koong, Prajnan Das, Vittorio Cristini, Ethan B. Ludmir, Eugene J. Koay

**Author notes:** **Corresponding authors:** Eugene J. Koay, MD PhD and Ethan B. Ludmir MD, Department of Gastrointestinal Radiation Oncology, Division of Radiation Oncology, The University of Texas MD Anderson Cancer Center Houston, Texas USA. These authors contributed equally to this work (Brian De, Prashant Dogra, Mohamed Zaid). **Prior presentation:** Preliminary data were presented at ASTRO 2016 (Boston, MA). **Author contributions:** Conception or design of the work: De, Dogra, Zaid, Cristini, Ludmir, Koay Acquisition, analysis, or interpretation of data: De, Dogra, Zaid, Elganainy, Sun, Amer, Wang Drafting the manuscript: De, Dogra, Zaid, Ludmir, Koay Critical revision of the manuscript for important intellectual content: All authors Statistical analysis: De, Dogra, Zaid Obtained funding: All authors Administrative, technical, or material support: All authors Final approval of the version to be published: All authors. **Data Sharing:** Research data are stored in an institutional repository and may be shared upon reasonable request to the corresponding author within one year of publication. **Statement of Translational Relevance:** Escalated doses of radiation (RT) for intrahepatic cholangiocarcinoma (iCCA) are associated with substantially prolonged survival in patients with unresectable disease. Understanding the mechanisms underlying radiation resistance in iCCA may improve patient outcomes. We hypothesized that changes in enhancement on computed tomography (CT) scans of iCCA would be indicative of the physical mechanisms of RT dose response. We developed a CT-informed mathematical model of RT response that yields patient-specific parameters of tumor biology. Results indicated that a CT-derived model-based tumor growth rate parameter was associated with local control and overall survival in iCCA. Furthermore, simulations with varying RT doses and fractionations for non-responders using patient-specific model parameters revealed optimized RT approaches to potentially convert non-responders to responders. These findings demonstrate the clinical utility of this CT-informed mathematical model to improve outcomes for iCCA, and may be applicable to other solid tumors for which changes in enhancement indicate a cytotoxic response.

## Abstract

**Background:** Although escalated doses of radiation therapy (RT) for intrahepatic cholangiocarcinoma (iCCA) are associated with durable local control (LC) and prolonged survival, uncertainties persist regarding personalized RT based on biological factors. Compounding this knowledge gap, the assessment of RT response using traditional size-based criteria via computed tomography (CT) imaging correlates poorly with outcomes. We hypothesized that quantitative measures of enhancement would more accurately predict clinical outcomes than size-based assessment alone and developed a model to optimize RT.

**Methods:** Pre-RT and post-RT CT scans of 154 patients with iCCA were analyzed retrospectively for measurements of tumor dimensions (for RECIST) and viable tumor volume using quantitative European Association for Study of Liver (qEASL) measurements. Binary classification and survival analyses were performed to evaluate the ability of qEASL to predict treatment outcomes, and mathematical modeling was performed to identify the mechanistic determinants of treatment outcomes and to predict optimal RT protocols.

**Results:** Multivariable analysis accounting for traditional prognostic covariates revealed that percentage change in viable volume following RT was significantly associated with OS, outperforming stratification by RECIST. Binary classification identified ≥33% decrease in viable volume to optimally correspond to response to RT. The model-derived, patient-specific tumor enhancement growth rate emerged as the dominant mechanistic determinant of treatment outcome and yielded high accuracy of patient stratification (80.5%), strongly correlating with the qEASL-based classifier.

**Conclusion:** Following RT for iCCA, changes in viable volume outperformed radiographic size-based assessment using RECIST for OS prediction. CT-derived tumor-specific mathematical parameters may help optimize RT for resistant tumors.

## Introduction

Intrahepatic cholangiocarcinoma (iCCA) is an uncommon and aggressive malignancy arising from the epithelial lining of the intrahepatic biliary tree, for which 70% of patients present with unresectable tumors. (1) While chemotherapy has historically been considered the standard of care for these patients, treatment with chemotherapy alone has been associated with unsatisfactory results, with median overall survival (OS) estimates ranging from 3 to 12 months. (2) Recent retrospective studies have shown that treatment with ablative doses of radiotherapy (RT) is associated with local control (LC) and OS. (3-5) However, the RT dose and fractionation schedule are subjectively chosen for each patient, and there is no consensus on the optimal protocol for treating iCCA. (3,6) Further compounding this uncertainty, evidence suggests that the optimal treatment of iCCA may be specific to each individual patient, potentially mediated by differences in pretreatments received, functional liver reserve, and intrinsic radiosensitivity. (7)

To address these intricacies, investigators in the field of mathematical oncology (8) have developed several mathematical models to predict radiotherapy response (9), with the widely used linear quadratic (LQ) model being the most prevalent. The LQ model posits that radiation-driven cell death results from double-strand DNA breaks caused directly by ionizing radiation, or through interactions involving single-strand DNA breaks. (10) Leder et al. formulated a more comprehensive model encompassing tumor cellular diversity and dynamically evolving radioresistance, enabling prediction of the efficacy of radiation schedules. (11) Prokopiou et al. introduced the proliferation saturation index (PSI), which implicitly links the proliferative fraction of a tumor with its radiation sensitivity. (12) Zahid et al. adapted the PSI model to simulate the impact of radiation not through LQ cell survival but by altering the tumor microenvironment, thereby enabling the forecasting of patient-specific responses to radiation. (13) To quantify radiosensitivity from a genomic perspective, modeling has been applied to develop a radiation sensitivity index (RSI) which connects the expression of specific genes to the radiosensitivity of cells. (14) Building on this, a genome-based model for adjusting radiotherapy dose (GARD) was developed by Scott et al., linking patient-specific RSI values and the LQ model with radiation dose plans to customize treatment schedules based on patient radiosensitivity. (15) To enhance the clinical application of mathematical models, we previously integrated pre-treatment CT imaging with mechanistic models to predict clinical responses in pancreatic cancer. These models are instrumental in understanding cytotoxic response mechanisms and have the potential to inform therapeutic decisions. (16) CT-based imaging for iCCA prior to and following RT provides a rich source of data from which mathematical models may permit therapy selection and adaptation.

Nevertheless, there are shortcomings in the current methods of radiographic evaluation of hepatic tumors. Conventionally, assessment of solid tumor response to systemic and locoregional therapies is performed with diagnostic imaging studies, which are crucial for prompt identification of responders and non-responders to treatment. Early prediction of treatment response after RT in iCCA may substantially impact further management, particularly if other liver-directed or systemic therapies are viable options. Currently, approved criteria for assessing tumor response to treatment include those from the World Health Organization (WHO) and Response Evaluation Criteria in Solid Tumors (RECIST). (17-19) Both criteria depend on changes in overall tumor size using the largest unidimensional or bidimensional measurements. A significant limitation of these size-based criteria is that they do not account for tumor morphological changes that reflect treatment response, such as tumor necrosis and decreased tumor perfusion, which may occur after liver-directed RT. (20)

To address this, an expert panel introduced the European Association for the Study of the Liver (EASL) criteria for hepatocellular carcinoma (HCC), which considers the enhancing portion of the tumor as viable tissue using bidimensional measurements of the largest enhancing portion of the tumor. (21) The EASL criteria have been shown to be predictive of patient outcomes and superior to RECIST and WHO criteria in the assessment of response after treatment of HCC. (22,23) Several studies have shown that three-dimensional quantitative EASL (qEASL) is predictive of survival after transarterial chemoembolization (TACE) and Yttrium-90 (Y-90) radioembolization in patients with HCC. (24,25) The utility of CT-based tumor enhancement for response assessment following RT for iCCA is yet to be explored and may provide more accurate outcome prediction than traditional size-based criteria. (26) More importantly, based on the dynamic assessment of viable tumor volume, adaptive radiotherapy can be supported by integration with mechanistic mathematical models of tumor growth and treatment response. (27) Therefore, the use of qEASL-based tumor enhancement measurements to develop mathematical models may be an attractive means of personalizing RT treatment protocols.

In the present study, we tested the hypothesis that changes in tumor enhancement following RT could inform a mathematical model of cytotoxic response in iCCA. We aimed to evaluate the utility of changes in tumor enhancement for RT response assessment in iCCA and predict outcomes following treatment. In addition, using qEASL-based viable tumor volume measurements, we sought to develop a mechanistic mathematical model as a prospective tool for personalization and adaptation of RT. An imaging-based biomarker of treatment response, integrated with mathematical modeling, may better guide the subsequent management of iCCA patients, which is highly important in both direct patient care and clinical trial settings.

## Methods

### Patient selection

We retrospectively identified 154 patients with biopsy-proven unresectable iCCA treated with definitive RT at MD Anderson Cancer Center from 2001-2021 and underwent liver-protocol computed tomography (CT) scans with non-contrast and portal venous phases before and after RT. CT imaging was deemed adequate for analysis if liver-protocol scans with and without contrast were available prior to the start of treatment and following treatment at the first follow-up visit. All patients were treated with 3D conformal radiotherapy, intensity-modulated photon radiotherapy (IMRT), conformal passive scatter proton beam, or intensity-modulated proton radiotherapy (IMPT) techniques. All patients had clinical and radiographic follow-up information. OS was measured from the initiation of RT until death or last follow-up. LC was calculated from the start of RT until local tumor progression or last follow-up. Local tumor progression was defined as any new tumor growth within or at the margin of the RT field. Imaging studies that revealed local tumor progression were reviewed and confirmed by a radiologist. The Institutional Review Board of MD Anderson Cancer reviewed the study under protocol PA14-0646 and waived the need for consent.

### Imaging selection and image analysis

All patients underwent pre- and post-RT (i.e., at the first follow-up visit) contrast-enhanced liver protocol CT imaging. We performed image visualization, processing, and analysis on the portal venous phase images of both CT scans using Philips IntelliSpace Portal 8 software (Philips Healthcare, Amsterdam, Netherlands). As shown in Figure 1A-D, after image registration, the corresponding non-contrast scan was subtracted from each post-contrast study to remove any artifactual enhancement (e.g., due to tumor calcification, biliary stents, or other metal artifact). The tumor volume was manually contoured on each 5 mm slice with exclusion of visually identifiable arteries or veins to reduce the possibility of contrast within larger vessels confounding measurements of tumor enhancement. Given that the degree of enhancement is a surrogate for tumor viability, inclusion of visually apparent vessels within the tumor contour could potentially lead to an artifactually increased estimate of viable tumors. (28)

**Figure 1.**
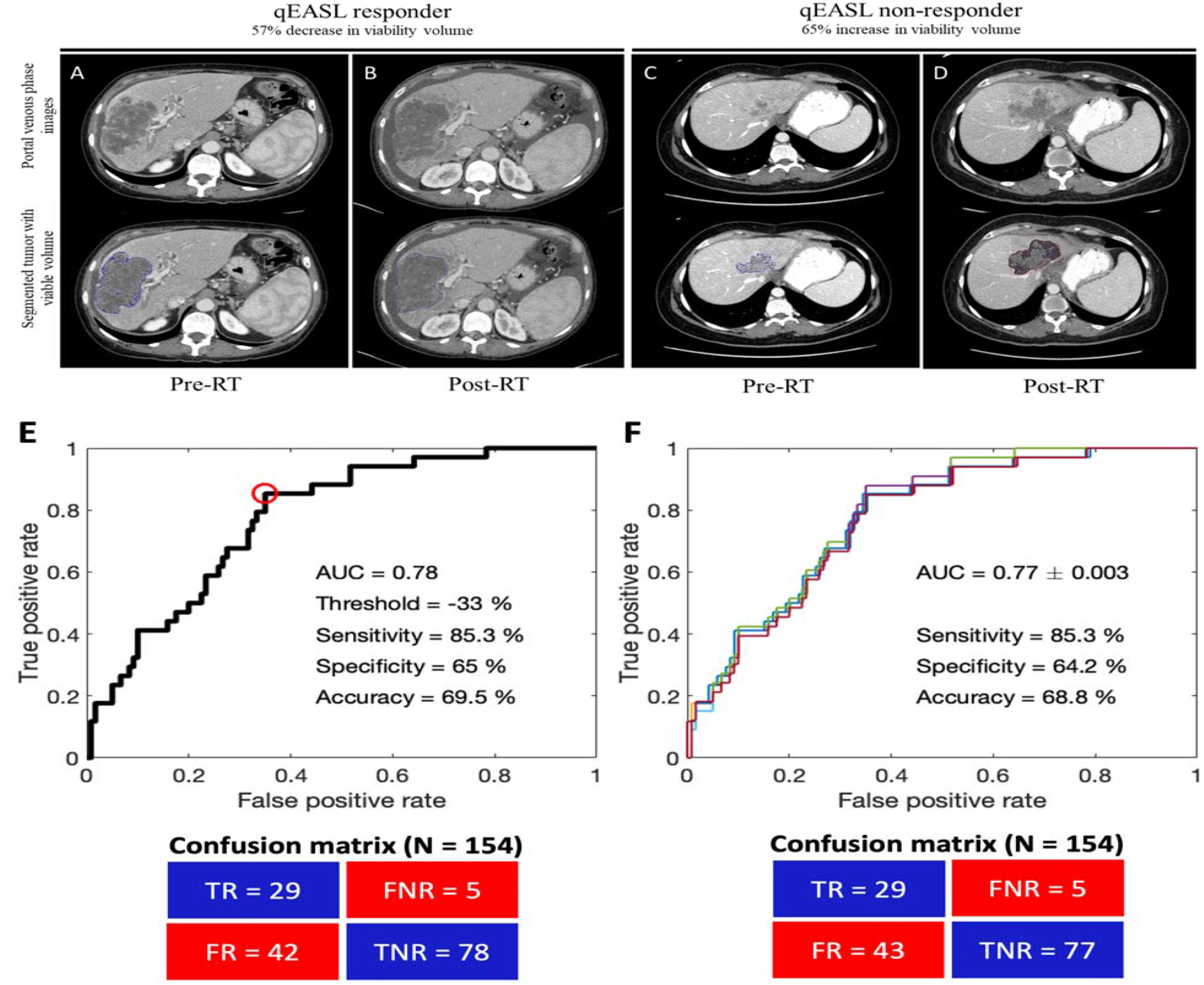
Binary classification of qEASL-based classifier. Representative CT images from A,C) pre-RT and B,D) post-RT in patients who were identified as A,B) a qEASL responder and C,D) a qEASL non-responder. E) ROC curve to determine qEASL-based cutoff point for percentage change in enhancement volume following RT. Red circle on the ROC curve denotes the optimal operating point identified through the Jaccard index. F) Cross validation of the binary classifier. The corresponding confusion matrices are displayed below the figures.

A reference region of non-tumor hepatic parenchyma was selected for each CT study. Viable tumor was classified as any tissue with enhancement exceeding one standard deviation of the mean enhancement measured in Hounsfield units from this reference region. Tumor contours and reference regions were independently verified by a radiation oncologist (EJK) who was blinded to all patient clinical data and outcomes.

### Molecular correlates of disease response

Mutation profiling was performed in 94 (61%) patients using previously described techniques, including next-generation sequencing from solid tumor tissue and/or circulating cell-free DNA to screen for single nucleotide variants, insertions/deletions, copy number gains, and gene fusions.(5) Associations between mutations and changes in enhancement were evaluated.

### Mathematical model development and calibration

To understand the mechanistic underpinnings of tumor response to RT in iCCA, we adapted our prior work to develop a mathematical model of tumor growth based on qEASL-derived measurements of viable tumor volumes. (16,29) The model (**Eq. 1**) describes the time-dependent evolution of the qEASL-derived tumor enhancement volume by accounting for exponential tumor growth and radiotherapy-induced tumor death. The model assumes that tumor enhancement volume kinetics is a surrogate for tumor growth kinetics, such that the rate of change of enhancement volume is given by

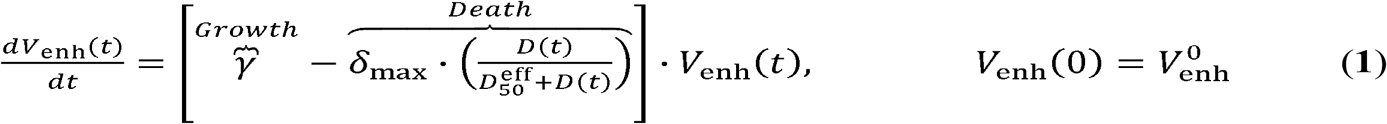

where *V*_enh_ (*t*) is the tumor enhancement volume at time *t, γ* is the first-order tumor growth rate constant, *δ*_max_ is the maximum tumor death rate constant (achievable under ideal conditions of radiation dose delivery and radiation sensitivity),*D*(*t*) is the radiation dose absorbed in the tumor at time *t*,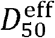 is the ‘effective’ half-maximal inhibitory dose of RT (i.e., potency of radiation), and 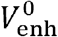 is the pre-treatment tumor enhancement volume designated as the initial condition. Note that the radiation-induced tumor death rate is proportional to the absorbed radiation dose *D*(*t*) and is assumed to be saturable; hence, the Michaelis-Menten function 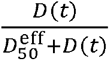 was used to model the death process.

We hypothesized that the potency of radiotherapy 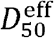depends on tumor perfusion (i.e., tissue oxygenation) (30) and non-specific molecular factors (e.g., activation of transcription factors to induce anti-apoptotic genes and upregulate cell proliferation) (31). Therefore, the ‘theoretical’ or *in vitro D*_50_ of radiotherapy is corrected for by a penalty factor obtained from the patient-specific tumor blood volume fraction (BVF), such that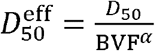. Here, BVF is a dimensionless quantity estimated as the ratio of tumor enhancement volume to total tumor exponent volume (pre-treatment); hence, BVF always assumes a value between 0 and 1. Here, the *α* is defined as the radiation resistance coefficient that represents the effect of patient-specific, ‘unknown’ molecular factors on radiation resistance. Therefore, a larger value of*α*leads to a smaller value of the denominator, thereby increasing 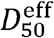 (i.e., reducing potency). Given the administered radiation dose per fraction(*D*_0_), it was modeled to undergo first-order decay such that the remaining total radiation in the tumor at time *t* was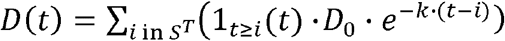. Here, *k* is the decay rate constant of radiation, and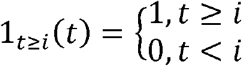is the indicator function, such that *i* represents individual treatment times, and *S*^*T*^ is the set of treatment times.

While some of the model parameters (Table S1) were either fixed based on average estimates from the literature (i.e., *D*_50_,*k,δ*_max_) (32,33) or calculated for individual patients from the available clinical data (i.e.,BVF from CT images), the remaining parameters were estimated by fitting the model to patient-specific tumor enhancement volume kinetics data (i.e.,*γ* and *α*;). The model was solved numerically as an initial value problem in MATLAB R2018a (MathWorks, Massachusetts, USA) using the built-in function *ode45* and nonlinear least-squares fitting of the model to clinical data was performed using the built-in function *lsqcurvefit*.

### Binary classification and cross-validation

Binary classifications were performed to obtain thresholds for i) qEASL-derived measures for treatment evaluation and ii) model-derived parameters (*γ*) for outcome prediction. In both cases, we used the entire dataset (n = 154) to train the binary classifiers. A logistic regression model was fitted between the predictor (i.e., percent change in enhanced volume for the first classifier and growth rate constant *γ* for the second classifier) and response variables (i.e., LC-based assessment of treatment outcome) using the built-in MATLAB function *fitglm*, and a receiver operating characteristic (ROC) curve was computed with the built-in MATLAB function *perfcurve*. Subsequently, using the Jaccard index(*J*) as a metric for classifier performance, the optimal operating point (or cutoff point) on the ROC curve was obtained for the two classifiers. Note that *J* can be obtained from a confusion matrix as *J=* TP/(TP+FP+FN),where TP, FP, and FN represent true positive, false positive, and false negative cases, respectively, and the operating point that yields the highest value of *J* is chosen as the optimal operating point. Based on the optimal operating point, the performance of the classifier was assessed from the confusion matrices by measuring the sensitivity (TP/(TP + FN)), specificity (TN/(TN + FP)), and accuracy ((TP + TN)/(TP + TN + FP + FN)), where TN represents true negatives.

The leave-one-out cross-validation (LOOCV) technique was used to validate the predictive ability of the binary classifiers. (34) For this, n-1 training datasets were generated from the total n data points by iteratively removing one data point. Each training dataset was used to generate a new ROC curve and to select a cutoff point (based on the calculations of *J*) to classify the left-out test data point. The prediction results from all the iterations were pooled to calculate the overall sensitivity, specificity, and accuracy of the classifiers. Note that these two classifications are mutually exclusive.

### Statistical analysis

Statistical analysis was performed using JMP Pro 16 (SAS Institute Inc., Cary, NC, USA) and Stata/SE 17.0 (StataCorp, College Station, TX, USA) software. Fisher’s exact test was used to compare the distributions of categorical variables between patient groups. Survival curves were constructed using the Kaplan-Meier method. The proportional hazards assumption was evaluated using tests of Schoenfeld residuals; all tests yielded *P*>0.05; thus, we failed to reject the null hypotheses that hazards were proportional. Cox proportional hazards models were used for univariate and multivariable survival analyses. Welch’s *t*-test (due to unequal sample sizes) was performed to evaluate differences in sample means for the modeling-related parameter values. A *P*-value ≤0.05 was considered significant for all analyses. To further test the predictive power of viability volume features, we used it to classify long-term (>24 months) versus short-term survival (≤24 months).

## Results

### Patient characteristics

Patient characteristics are shown in Table 1. The analytic cohort consisted of 154 patients. The median age was 64 years (range, 29-88 years), and 74 (48%) patients were female. The median RT dose was 62.5 Gy (range, 35-100 Gy), and the median number of fractions was 15 (range, 3-31 fractions). The median interval between the end of RT and post-RT CT scan was 1.8 months (range, 0.1-10.9 months). RT technique was proton in 37 patients and photon in 117 patients based on physician preference.

**Table 1.** Patient characteristics among all patients and stratified by qEASL response.

|  | All patients | qEASL responders | qEASL non-responders | P-value |
| --- | --- | --- | --- | --- |
| **Number** | 154 | 72 | 82 |  |
| **Age, median [range]** | 64 [29-88] | 63 [29-86] | 65 [33-88] | 0.069 |
| **Sex** |  |  |  |  |
| Female | 74 | 36 | 38 | 0.747 |
| Male | 80 | 36 | 44 |  |
| **Time interval between end of RT and post-RT CT scan (months), median [range]** | 1.8 [0.1-10.9] | 1.9 [0.2-9.3] | 1.8 [0.1-10.9] | 0.051 |
| **AJCC 8<sup>th</sup> edition stage** |  |  |  |  |
| I | 12 | 7 | 5 | 0.385 |
| II | 29 | 16 | 13 |  |
| III | 20 | 6 | 14 |  |
| IVA | 44 | 22 | 22 |  |
| IVB | 49 | 21 | 28 |  |
| **Tumor largest dimension (mm), median [range]** | 72 [17-179] | 74 [17-179] | 72 [24-176] | 0.865 |
| **Intrahepatic satellites at diagnosis** |  |  |  |  |
| Yes | 76 | 31 | 45 | 0.202 |
| No | 78 | 41 | 37 |  |
| **ECOG scale** |  |  |  |  |
| 0 | 52 | 22 | 30 | 0.687 |
| 1 | 90 | 44 | 46 |  |
| 2 | 11 | 6 | 5 |  |
| 3 | 1 | 0 | 1 |  |
| **Concurrent chemotherapy** |  |  |  |  |
| No | 52 | 28 | 24 | 0.234 |
| Yes | 102 | 44 | 58 |  |
| **Radiation dose Gy median [range]** | 62.5 [35-100] | 66 [50-100] | 60 [35-100] | 0.124 |
| **BED ≥ 80.5** |  |  |  |  |
| Yes | 113 | 58 | 55 | 0.069 |
| No | 41 | 14 | 27 |  |
| **Technique** |  |  |  |  |
| Photon | 117 | 47 | 70 | 0.004* |
| Proton | 37 | 25 | 12 |  |

\* Significant at 5% level. Abbreviations: AJCC: American Joint Committee on Cancer; BED: Biologically effective dose; CT: Computed tomography; Gy: Gray; ECOG: Eastern Cooperative Oncology Group; qEASL, Quantitative European Association for the Study of Liver Criteria

### Disease outcomes

Patient outcomes following RT have been previously reported by our institution. (3,5) In the current cohort, updated with the last available follow-up, the median follow-up time was 64 months (range, 43-80 months) following RT. At latest follow up, 63 patients (41%) had experienced local failure and 127 (82%) had died. The median LC was 30 months (95% confidence interval [CI] 19-44 months), and median OS was 18 months (95% CI, 15-21 months).

### RECIST 1.1 assessment

Patients were classified by RECIST 1.1 criteria as having complete response (CR; no visible tumor), partial response (PR; decrease of ≥30% in sum of longest diameter), progressive disease (PD; increase of ≥20% in sum of longest diameter), or stable disease (SD; all others). (19) At the first follow up scan after completion of RT, the median longest tumor diameter did not significantly change; the median was 7.2 cm (interquartile range [IQR], 5.2-10.3 cm) before RT and 6.8 cm (IQR, 4.9-10.0 cm; *P*=0.115) after RT. Utilizing the RECIST 1.1 criteria, the training set had 8 patients with PR to RT, 86 patients with SD, 14 patients with PD, and no patients with CR. As shown in Figure 2A-B, there was no significant difference in OS between the various response categories (*P*=0.7052), but there was a significant difference in LC (*P*=0.0137) when patients were stratified by RECIST 1.1 response.

**Figure 2.**
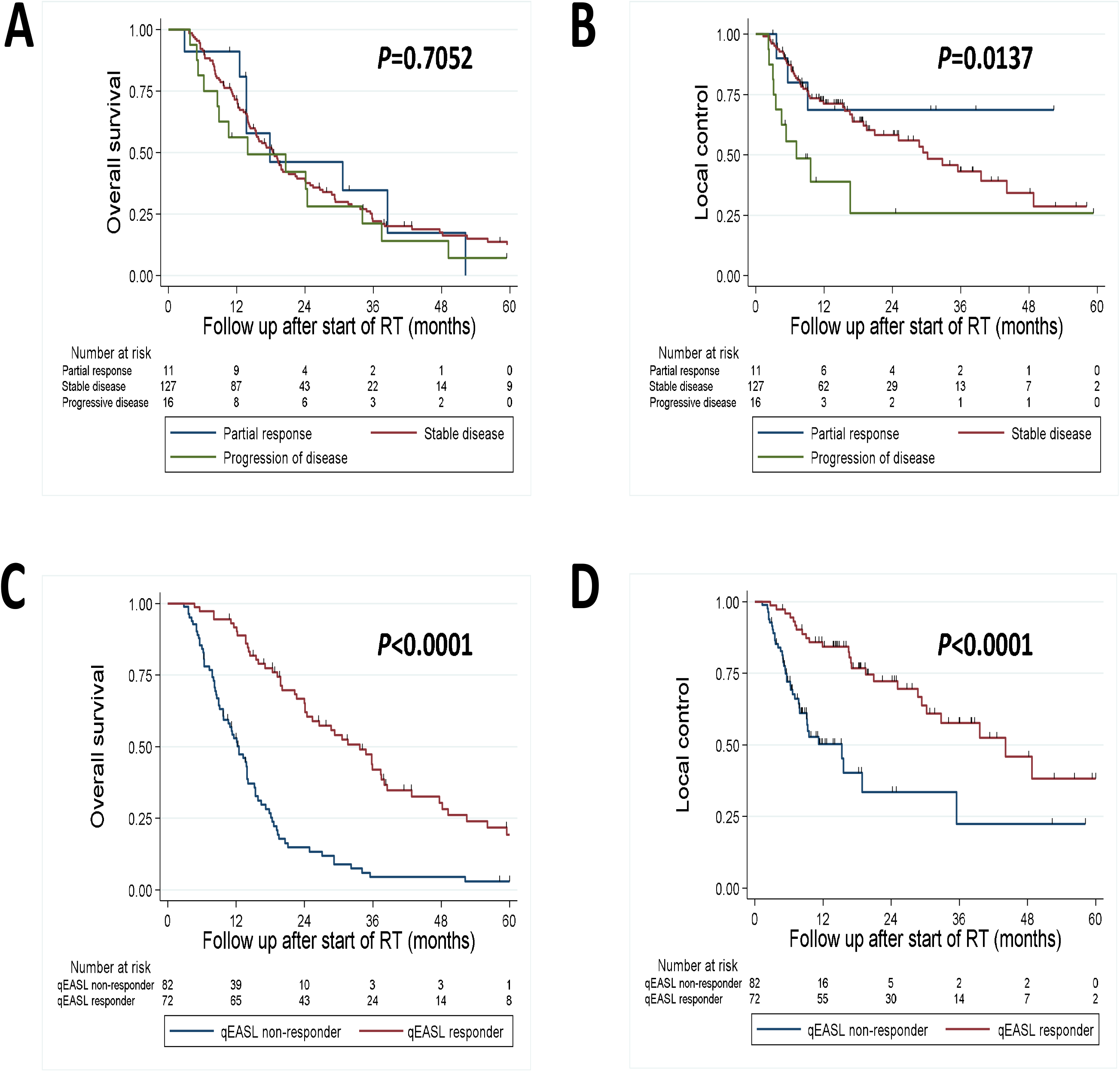
Comparison of survival curves stratified by RECIST and qEASL-based evaluations. Overall survival (A) and local control (B) among patients stratified by response as measured by RECIST 1.1 and overall survival (C) and local control (D) among patients stratified by response as measured by qEASL.

### qEASL assessment

Baseline characteristics stratified by qEASL (responders and non-responders) are shown in Table 1. A key difference was greater use of proton therapy in the qEASL-responder cohort. As shown in Table S2, univariate Cox analysis showed that a decrease in qEASL-derived viable volume was associated with prolonged OS (*P*<0.001) but not LC (*P*=0.083). Similarly, in multivariable analysis, a decrease in viable volume as a continuous variable was significantly associated with longer OS (*P*<0.001) but not LC (*P*=0.088) after adjusting for age, stage, and BED. The change in viable volume (%) was evaluated for its ability to distinguish responders from non-responders.

Using binary classification, a cut-off point of 33% decrease in viable volume was determined to optimally predict prolonged LC (>24 months), i.e., being a responder (Figure 1E; area under the ROC curve (AUC) = 0.78). The corresponding sensitivity, specificity, and accuracy of the classifier were 85.3%, 65.0%, and 69.5%, respectively. Further, as shown in Figure 1F, LOOCV upheld the validity of the classifier with an average AUC of 0.77±0.003 and sensitivity, specificity, and accuracy of predicting the left-out data point at 85.3%, 64.2%, and 68.8%, respectively. Furthermore, patients classified as responders according to the qEASL criteria demonstrated a significantly longer median OS (34 vs. 13 months; *P*<0.0001) and median LC (44 vs. 15 months; *P*<0.0001) (Figure 2C-D).

### Molecular correlates of response to treatment

Mutational frequencies and their associations with treatment responses are shown in Table S3. The most commonly observed mutations were in *TP53, IDH1, BAP1*, and *ARID1A*. Univariate linear regression was performed to identify potential relationships between mutations and treatment response, as measured by the percentage change in enhancement volume following RT. Only one potential association was identified: *ARID1A* mutation was associated with a 281% *increase* in enhancement volume vs. a 13% *decrease* in enhancement volume in *ARID1A* wild type patients (*P*=0.034).

### Modeling-derived predictor of treatment response

As shown in Figure 3A,B, the model-fitted trajectory of patient tumor enhancement volume over time (orange line), under the patient-specific radiation treatment implemented in the simulation (blue line), demonstrates the difference in the evolution of tumor dynamics in a representative responder (patient ID 54; Figure 3A) and non-responder (patient ID 47; Figure 3B). The key model parameter values, estimated from CT imaging (i.e., BVF) and model fitting (i.e.,*γ* and *α*), shown in the inset of the figures, suggest that in the specific examples, despite the same dose and fractions given to the two patients, the tumor response varied significantly owing to differences in tumor growth rate *γ* and BVF. A responder has a slower growth rate and higher BVF than a non-responder. As we know from the model equation, because of the difference in BVF, the 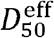 in the two cases varied by an order of magnitude, such that the potency of radiation in the non-responder appears to be much lower than that in a responder. In addition, a larger value of *γ* obtained for a non-responder suggests a more aggressively growing tumor, indicating a net positive growth rate under treatment.

**Figure 3.**
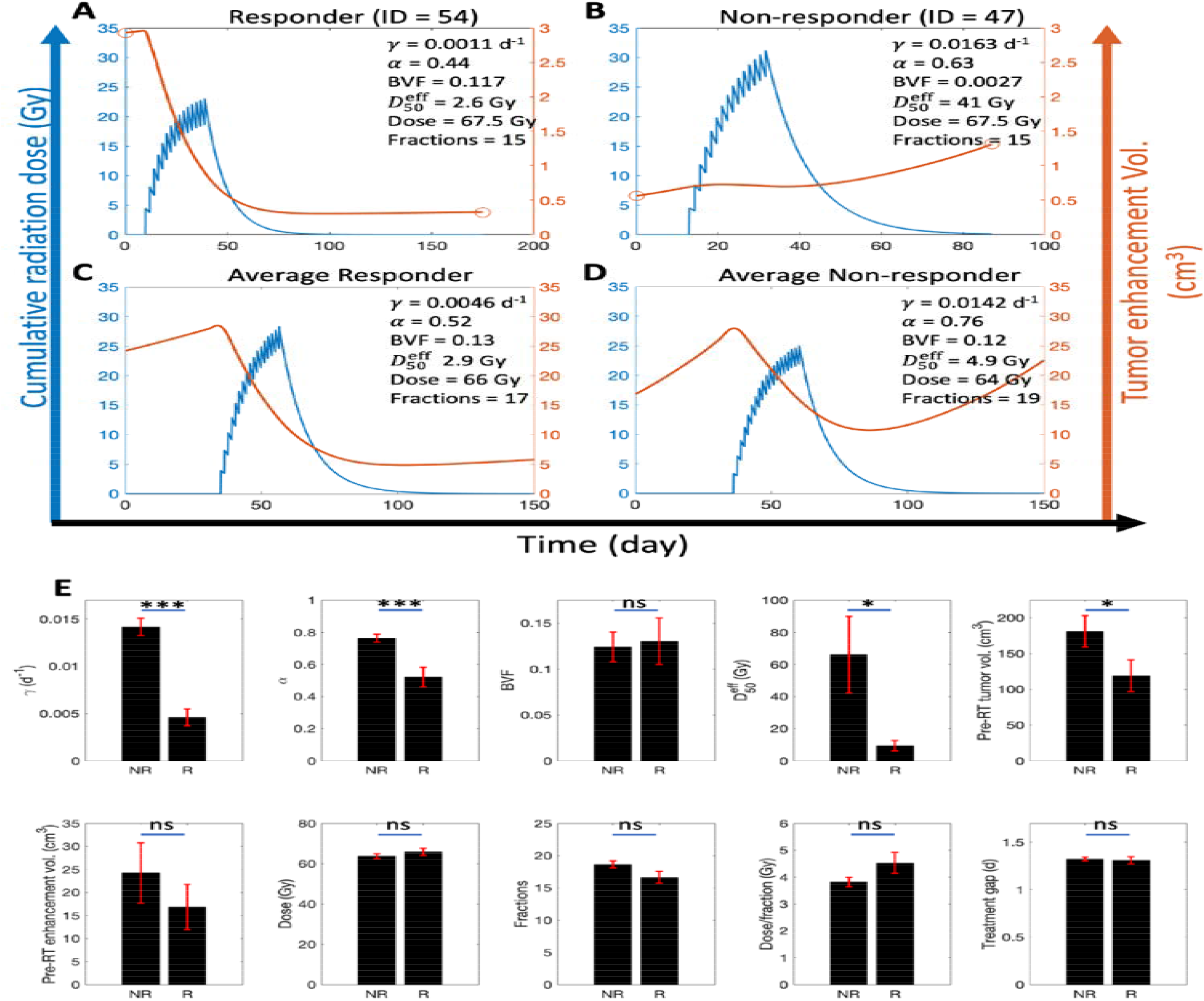
Representative model simulations and parameter estimates. Model fits to tumor enhancement volume data (orange) are shown, along with the simulated, patient-specific radiation treatment cycles (blue) for a representative A) responder (patient ID 54) B) non-responder (patient ID 47), C) average responder, and E) average non-responder subject. Orange circles denote the raw data for enhancement volumes obtained from CT images. Parameter estimates for the corresponding cases are given in the inset. E) Model and data-derived parameter estimates. The height of each bar represents the mean (n=154) and error bars represent standard error of the mean. Welch’s *t*-test was used to identify differences between responders (R) and non-responders (NR). * *P*< 0.05; *** *P*< 0.0001; ns: non-significant. Note that treatment gap (days) represents the average time interval between two fractions.

Furthermore, the model was fit individually to every patient (Figure S1), and parameter estimates were obtained (Figure 3E, Table S5). Using the average parameter values shown in Figure 3E, we simulated the tumor response dynamics for an average responder and an average non-responder, starting with the average initial tumor enhancement volume across all patients, that is, ∼25 cm^3^ and ∼17 cm^3^, respectively (Figure 3C-D). From these simulations, we observed that the average responder showed a ∼51% reduction in enhancement, whereas the average non-responder exhibited only ∼11% decrease in enhancement during the course of treatment. Extending the simulation beyond the end of treatment up to day 150 demonstrated that as the dose washes out, tumor enhancement eventually begins to increase, although at much different rates for the two groups (due to significant differences in their *γ* parameter values), such that the enhancement volumes at day 150 were 6 cm^3^ and 22.5 cm^3^ for responders and non-responders, respectively.

The above observations can be contextualized from the parameter estimates shown in Figure 3E. The tumor growth rate constant *γ*, radiation resistance coefficient *α*, effective half-maximal inhibitory radiation dose or radiation potency 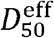, and pre-RT viable volume were the only parameters that were significantly different between the responder and non-responder groups (Welch’s *t*-test, *P*<0.05). This indicates that owing to relatively smaller growth rates, lesser radiation resistance, and higher radiation potency, the responder group showed better treatment outcomes to the standard RT regimens. Note that while the pre-RT tumor volumes were significantly smaller in the responder cohort, the corresponding difference between the BVF values (which is the ratio of pre-RT tumor enhancement volume to pre-RT tumor volume) between the two groups was insignificant. This suggests that 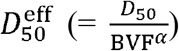 is predominantly affected by the desensitization coefficient *α*.

We further tested the four parameters identified as significantly different between the responder and non-responder groups for their ability to predict treatment outcomes by performing logistic regression-based binary classification. As shown in Figure 4A, the ROC curves obtained for the four classifiers (*α, β*,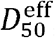, and pre-RT tumor volume) had AUC values of 0.71, 0.81, 0.69, and 0.57, respectively. Based on the obtained AUC values, we proceeded with growth rate *γ* as a potential model-derived predictor of treatment outcomes and calculated the optimal operating point on the ROC curve using the Jaccard index (Figure 4B). From this analysis, 0.0042 d^-1^ (doubling time of 165 d) was identified as the cutoff value to differentiate responders from non-responders, such that patients with estimated tumor growth rates≤0.0042 d^-1^ were more likely to be responders, while those with growth rates >0.0042 d^-1^ were more likely to be non-responders. Although the ability of *γ* to correctly identify true responders was somewhat low (sensitivity = 61.8%), its ability to correctly identify true non-responders was quite high (specificity = 85.8%), leading to an overall prediction accuracy of 80.5%. The results were validated using the LOOCV method, which led to an average AUC value of 0.816 ± 0.0032 calculated across the 154 ROC curves obtained by retraining the classifier 154 times upon removing a new data point every time (Figure 4C). The ability of the classifier to correctly predict the left-out data point during LOOCV led to a specificity of 83.3%, sensitivity of 52.9%, and an overall accuracy of 76.6%. The corresponding confusion matrices are shown in Figure S2.

**Figure 4.**
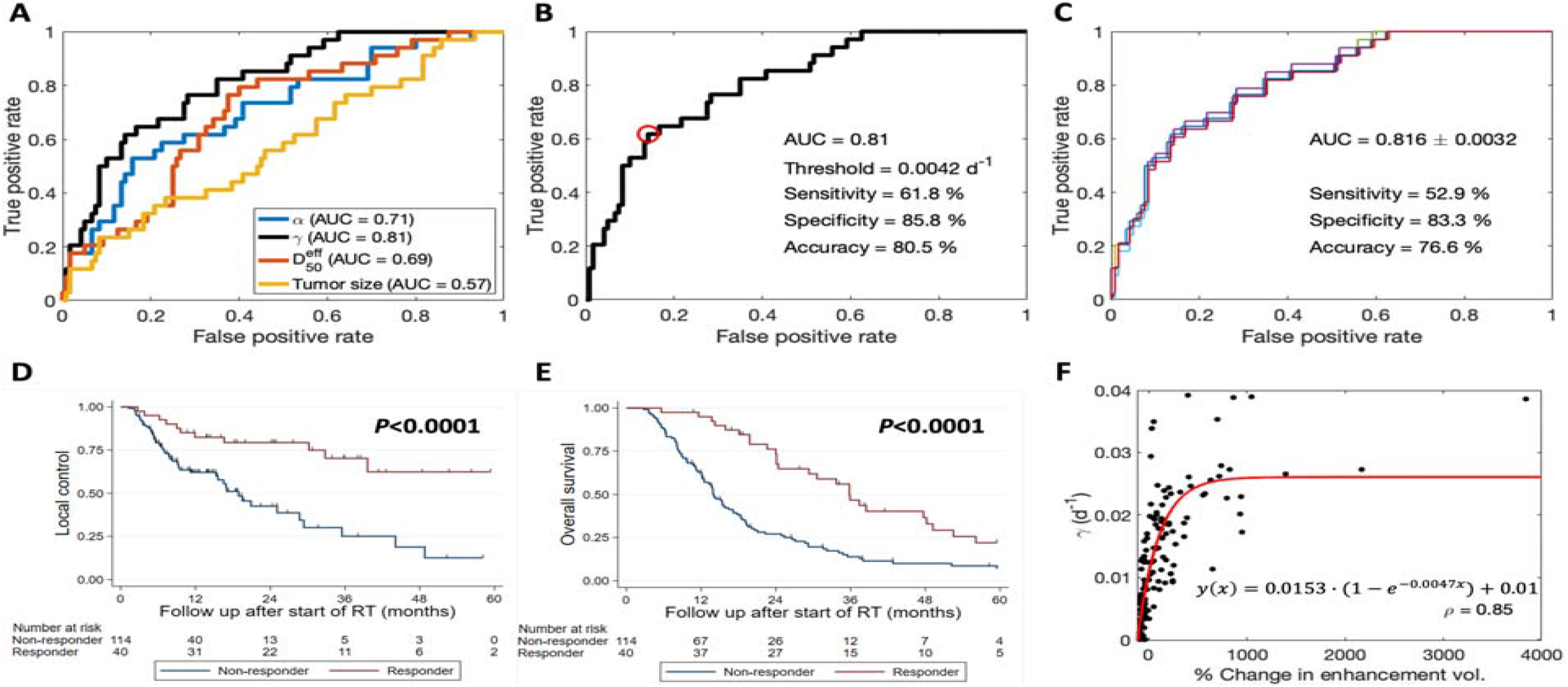
Binary classification of mathematical model-based classifier. A) ROC curves corresponding to *α,γ*, 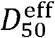, and pre-RT tumor volume as binary classifiers. The associated AUC values of the curves are given in the legend. B) ROC curve corresponding to the y classifier, highlighting the optimal operating point (red circle) and performance metrics (in the inset). C) LOOCV of the *γ* classifier showing 154 ROC curves. The corresponding confusion matrices are displayed in Figure S2. Survival curves stratified by qEASL growth rate. Overall survival (D) and local control (E) among patients stratified by qEASL-based growth rate. F) Spearman’s correlation between model-derived predictor of response *(γ*) and qEASL-derived measure of treatment outcome assessment (% change in enhancement volume).

The classifier was further tested for its ability to predict survival using Kaplan-Meier survival curves, log-rank test, and Cox analysis. The survival curves for OS and LC stratified by the growth rate-based binary classifier are shown in Figure 4D-E. A clear separation was observed between the two cohorts for both OS and LC (*P*<0.001 for both). The median OS for responders and non-responders was 36 months (95% CI 24-48) and 14 months (95% CI 12-17), respectively. The median LC for non-responders was 19 months (95% CI 15-29) and the median LC for responders was not reached at last follow up. In univariate Cox analysis, compared to as a non-responders, classification as a responder was associated with an HR of 0.40 (95% CI 0.26-0.61; *P*<0.001) for death and a HR of 0.29 (95% CI 0.15-0.56; *P*<0.001) for local progression. (Table S4)

Finally, to establish the validity of the model-based *γ* classifier to work on the qEASL-based measure of treatment response or as a surrogate for the (%) change in the viable volume classifier, we assessed the correlation between model-derived *γ* and the percentage change in viable volume. As shown in Figure 4F, a strong monotonic correlation was observed between the two variables, indicated by Spearman’s rank correlation coefficient *ρ* = 0.85, and defined by the function *y*(*x*) =0.0153·(1−*e* ^−0.0047*·x*^). This suggests that the model-predicted *γ* values are likely to correctly classify the patient outcomes evaluated using the qEASL-based response measure.

### Model-based treatment optimization

As shown in Figure 5, the model was tested as a means to optimize or personalize the treatment regimen to improve outcomes. In Figure 5A,B, the dashed black lines represent the tumor enhancement volume kinetics of the average responders and non-responders examined in Figures 3C,D. The model was used to simulate different clinically relevant treatment protocols and to predict the corresponding tumor enhancement volume kinetics while keeping the model parameter values constant and equal to the values for the average subjects. As observed, for both responders and non-responders, the only treatment regimen that outperformed the average behavior was 100 Gy in 25 fractions (BED10 140 Gy). Simulations were conducted to assess the differential impact of latency to RT starting from the time of initial radiographic imaging by examining the predicted change in enhancement on day 60 following the conclusion of RT (Table S6). Specifically, the model predicts that starting RT seven days sooner may benefit patients who ultimately do not respond to RT.

**Figure 5.**
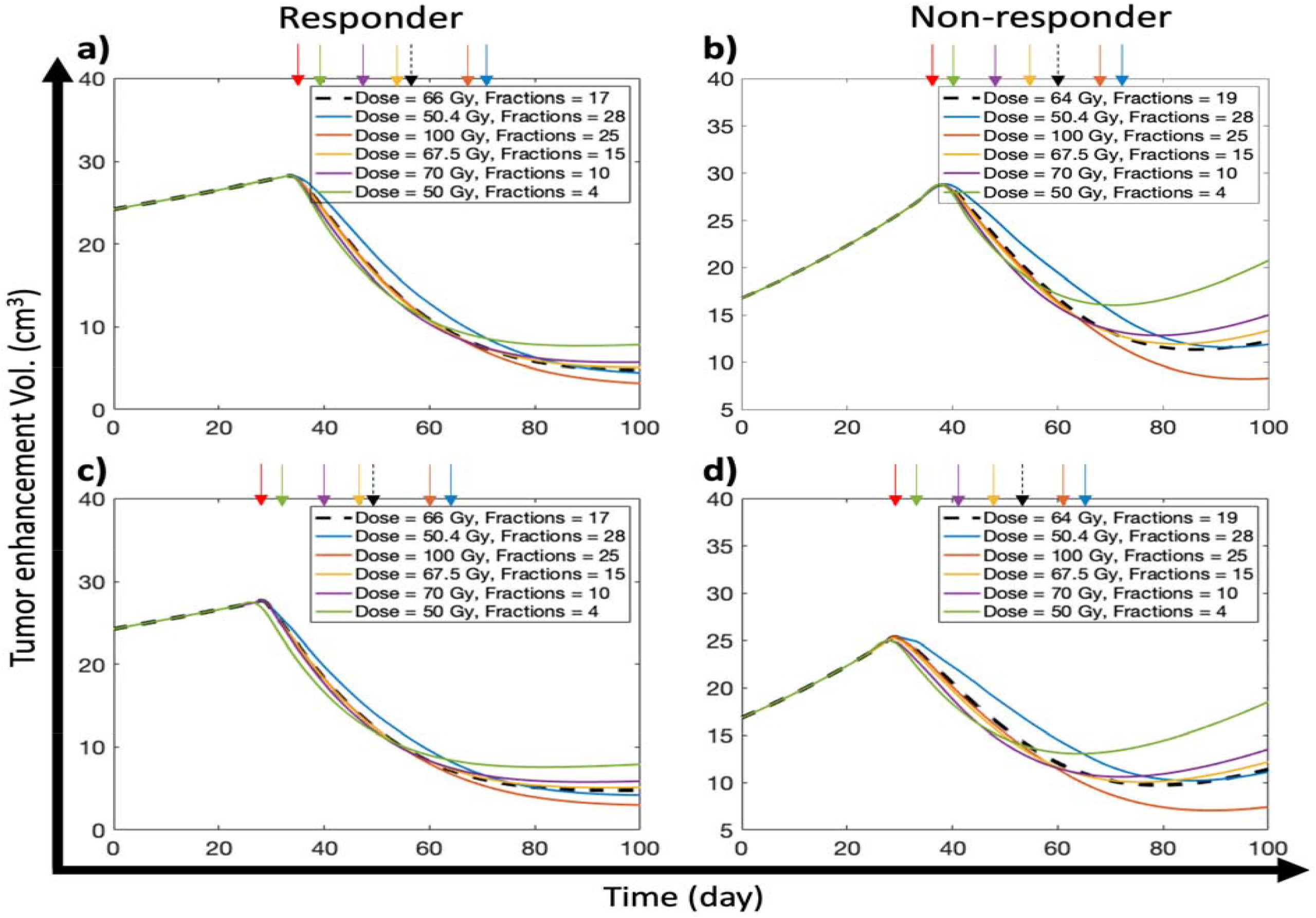
Treatment optimization simulations. Model-based predictions of tumor enhancement volume kinetics corresponding to a,c) the average responder and b,d) average non-responder subject under different treatment conditions. Dashed black lines represent the response kinetics corresponding to the average subject simulated in Figure 4c,d. In c,d) treatment was initiated seven days sooner than in a,b). Red arrow indicates the initiation time of treatment, and the other colored arrows denote the termination time of a given treatment (described in legend).

## Discussion

Early identification of responders and non-responders to RT for patients with iCCA has been elusive, owing in part to suboptimal prediction while using traditional size-based radiographic criteria. Without prompt assessment of the response to treatment, clinicians cannot make informed decisions about the most appropriate modifications to a patient’s treatment course. In the current study, we developed a mechanistic model using changes in tumor enhancement following RT to better understand tumor growth kinetics and accurately predict the response to treatment. We found that tumor viability, as estimated by the degree of post-contrast enhancement, outperformed the traditional size-based criteria and effectively stratified patients with respect to LC and OS following RT, independent of other clinical characteristics, treatment parameters, or prognostic features. Furthermore, mathematical modeling of viable tumor volumes revealed earlier initiation of RT as a potential strategy to overcome anticipated non-response to RT.

Personalization of RT dose and fractionation is a captivating potential application of the results of the current study. The developed mathematical model provides a mechanistic description of tumor response to radiation and helps identify a translatable outcome predictor for prospective clinical applications. The model accounts for tumor growth dynamics and radiation-induced tumor death, while considering the role of tumor perfusion (BVF) and other molecular factors *(α*) in radiation resistance. Since the model is built on qEASL-derived data for viable tumor volume, it may enable more accurate prediction of radiation response, given that the functional or viable volume of the tumor is a more realistic determinant of treatment response, unlike the gross size measured via RECIST, which did not predict outcomes as well as qEASL. Therefore, through characterization of tumor- and treatment-specific parameters from routine clinical measurements or population averages, our model can predict viable tumor volume trajectories, which can provide the basis for treatment design optimization or adaptation in real time for improved outcomes. Importantly, modeling-based analysis revealed the significance of tumor growth rates in determining treatment outcomes, as demonstrated by the high accuracy of *γ*(AUC of ROC = 0.81) in classifying patient responses. This suggests that a net positive growth rate is maintained by tumors of non-responders, which can perhaps be attributed to molecular factors that upregulate cell proliferation or induce anti-apoptotic genes. (31) This is consistent with the literature, where pre-treatment tumor growth rates in various solid tumors have been found to negatively affect treatment outcomes with immune checkpoint inhibitors (35,36) and chemoradiotherapy (37-39). We foresee the application of this finding to estimate tumor growth rate using pre-treatment imaging, thereby predicting treatment outcomes and allowing adaptation of treatment design for improved outcomes.

Furthermore, as a representative example, through model simulations, we demonstrated that longer fractionation schedules (25 fractions), combined with higher doses (100 Gy), appeared to be correlated with greater decreases in tumor enhancement, capable of transforming the average non-responder into a responder. These results are consistent with prior work showing a potential LC and OS benefit to ablative doses of RT. (3) We also show that the timing of RT may have a significant impact on change in enhancement following RT, particularly for the group of patients who were deemed to be non-responders. As a next step, identification of non-responders using pre-RT or even earlier CT-based features will be crucial in further investigating if a cohort of patients for whom more prompt treatment, longer fractionation, or higher doses may be warranted.

As shown in our study population, radiographic assessment using size-based criteria does not provide a complete picture of response to therapy. Consideration of post-treatment morphological changes is essential for a deeper understanding of treatment effects and the different patterns of tumor response. With tumor evaluation using RECIST, most patients in this study (82%) would have been categorized as “stable disease”, despite the wide variability in both tumor control and survival in this cohort. This highlights the relative insensitivity of size-based response criteria in predicting outcomes. Conversely, qEASL has been shown to have higher sensitivity and specificity in estimating tumor response than size-based assessments. (40) With the present analysis, we have now extended the findings of qEASL response to patients with iCCA treated with RT, supporting the idea that enhancement metrics in hepatobiliary cancers are linked to disease biology and treatment response. (40-45)

As an exploratory analysis, we reviewed molecularly characterized patients and identified two potential relationships between mutation status and changes in enhancement. Although the subgroup sample sizes were small, *ARID1A* mutation was associated with an increase in enhancement volume. Outcome data for patients with *ARID1A* mutations in biliary tract cancers are limited, but a few studies have suggested a poorer prognosis for these patients, which is consistent with the observed increases in enhancement. (46,47) Further data are needed to more robustly characterize the associations between molecular profile and treatment response.

We acknowledge that our study was limited by its retrospective nature and number of patients. This cohort spanned 20 years, during which time treatment strategies have evolved. As such, there is heterogeneity in radiation doses and techniques, chemotherapeutic agents, and criteria for patient selection for therapy, which may limit the generalizability of our findings. Nevertheless, the ability to measure enhancement over a long period of time despite the use of different treatment modalities is also a strength of the methodology, indicating that assessment by qEASL can be used over a variety of acquisition parameters and clinical practices. Our future directions include independent validation of the reported findings, improvement of tumor registration accuracy through the implementation of biomechanical-registration modeling, ascertainment of the impact of upfront systemic therapy, and investigation of the enhancement pattern mapping of these tumors. (48-50)

In summary, mathematical models utilizing tumor enhancement volume changes following RT may be used to assess response to therapy as early as the first follow-up scan. This measurement outperforms RECIST, which is the standard criterion used for assessing radiographic response. In addition, measurement of tumor growth rate pre-treatment can serve as a predictor of treatment outcomes, which, upon integration with the mathematical model, can be used for treatment personalization. Further validation of these findings using prospective data will help establish viable volume changes as an early signal of treatment response, which may better guide the subsequent management of iCCA patients treated with RT.

## Data Availability

The data that support the findings of this study are available from the corresponding author, EJK, upon reasonable request, and with respect to maintaining patient confidentiality.

## List of abbreviations

RT: radiotherapy
A-RT: ablative radiotherapy
iCCA: intrahepatic cholangiocarcinoma
RECIST: Response Evaluation Criteria in Solid Tumors
qEASL: Quantitative European Association for the Study of Liver criteria
LOOCV: = Leave-one-out cross validation
OS: overall survival
LPFS: local progression-free survival
World Health Organization(WHO): 1D = 1-dimensional, 2D = 2-dimensional, 3D = 3-dimensional, TACE transarterial chemoembolization
ROC: receiver operating characteristic
CR: complete response
PR: partial response
PD: progressive disease
SD: stable disease
ROC: receiver operating characteristic
AUC: area under the curve
BED: biologically effective dose

## Declarations

### Ethics approval and consent to participate

The Institutional Review Board Committee at MD Anderson Cancer Center approved the review of the medical records of these patients (PA14-0646). The need for informed patient consent was waived because this was a retrospective review, and no identifiable patient information was included in this report. This study was conducted in accordance with the ethical standards of the Declaration of Helsinki and its amendments.

### Consent for publication

Not applicable.

## Acknowledgments

Not applicable.

## Authors’ information

Not applicable.

## References

1. Vauthey JN, Blumgart LH. Recent advances in the management of cholangiocarcinomas. Seminars in liver disease 1994;14(2):109–14 doi 10.1055/s-2007-1007302.

2. Thongprasert S. The role of chemotherapy in cholangiocarcinoma. Annals of oncology : official journal of the European Society for Medical Oncology / ESMO 2005;16 Suppl 2:ii93–6 doi 10.1093/annonc/mdi712.

3. Tao R, Krishnan S, Bhosale PR, Javle MM, Aloia TA, Shroff RT, et al. Ablative Radiotherapy Doses Lead to a Substantial Prolongation of Survival in Patients With Inoperable Intrahepatic Cholangiocarcinoma: A Retrospective Dose Response Analysis. Journal of clinical oncology : official journal of the American Society of Clinical Oncology 2015 doi 10.1200/jco.2015.61.3778.

4. Smart AC, Goyal L, Horick N, Petkovska N, Zhu AX, Ferrone CR, et al. Hypofractionated Radiation Therapy for Unresectable/Locally Recurrent Intrahepatic Cholangiocarcinoma. Ann Surg Oncol 2020;27(4):1122–9 doi 10.1245/s10434-019-08142-9.

5. De B, Abu-Gheida I, Patel A, Ng SSW, Zaid M, Thunshelle CP, et al. Benchmarking Outcomes after Ablative Radiotherapy for Molecularly Characterized Intrahepatic Cholangiocarcinoma. J Pers Med 2021;11(12) doi 10.3390/jpm11121270.

6. De B, Tran Cao HS, Vauthey JN, Manzar GS, Corrigan KL, Raghav KP, et al. Ablative liver radiotherapy for unresected intrahepatic cholangiocarcinoma: Patterns of care and survival in the United States. Cancer 2022.

7. Normolle D, Pan C, Ben-Josef E, Lawrence T. Adaptive trial of personalized radiotherapy for intrahepatic cancer. Per Med 2010;7(2):197–204 doi 10.2217/pme.10.5.

8. Cristini V, Lowengrub J. Multiscale modeling of cancer: an integrated experimental and mathematical modeling approach. Cambridge University Press; 2010.

9. Bekker RA, Kim S, Pilon-Thomas S, Enderling H. Mathematical modeling of radiotherapy and its impact on tumor interactions with the immune system. Neoplasia 2022;28:100796 doi 10.1016/j.neo.2022.100796.

10. Chadwick KH, Leenhouts HP. A molecular theory of cell survival. Phys Med Biol 1973;18(1):78–87 doi 10.1088/0031-9155/18/1/007.

11. Leder K, Pitter K, LaPlant Q, Hambardzumyan D, Ross BD, Chan TA, et al. Mathematical modeling of PDGF-driven glioblastoma reveals optimized radiation dosing schedules. Cell 2014;156(3):603–16 doi 10.1016/j.cell.2013.12.029.

12. Prokopiou S, Moros EG, Poleszczuk J, Caudell J, Torres-Roca JF, Latifi K, et al. A proliferation saturation index to predict radiation response and personalize radiotherapy fractionation. Radiat Oncol 2015;10:159 doi 10.1186/s13014-015-0465-x.

13. Zahid MU, Mohsin N, Mohamed ASR, Caudell JJ, Harrison LB, Fuller CD, et al. Forecasting Individual Patient Response to Radiation Therapy in Head and Neck Cancer With a Dynamic Carrying Capacity Model. Int J Radiat Oncol Biol Phys 2021;111(3):693–704 doi 10.1016/j.ijrobp.2021.05.132.

14. Eschrich S, Zhang H, Zhao H, Boulware D, Lee JH, Bloom G, et al. Systems biology modeling of the radiation sensitivity network: a biomarker discovery platform. Int J Radiat Oncol Biol Phys 2009;75(2):497–505 doi 10.1016/j.ijrobp.2009.05.056.

15. Scott JG, Berglund A, Schell MJ, Mihaylov I, Fulp WJ, Yue B, et al. A genome-based model for adjusting radiotherapy dose (GARD): a retrospective, cohort-based study. Lancet Oncol 2017;18(2):202–11 doi 10.1016/s1470-2045(16)30648-9.

16. Wang CX, Elganainy D, Zaid MM, Butner JD, Agrawal A, Nizzero S, et al. Mass Transport Model of Radiation Response: Calibration and Application to Chemoradiation for Pancreatic Cancer. Int J Radiat Oncol Biol Phys 2022;114(1):163–72 doi 10.1016/j.ijrobp.2022.04.044.

17. Miller AB, Hoogstraten B, Staquet M, Winkler A. Reporting results of cancer treatment. Cancer 1981;47(1):207–14.

18. Therasse P, Arbuck SG, Eisenhauer EA, Wanders J, Kaplan RS, Rubinstein L, et al. New guidelines to evaluate the response to treatment in solid tumors. European Organization for Research and Treatment of Cancer, National Cancer Institute of the United States, National Cancer Institute of Canada. J Natl Cancer Inst 2000;92(3):205–16.

19. Eisenhauer EA, Therasse P, Bogaerts J, Schwartz LH, Sargent D, Ford R, et al. New response evaluation criteria in solid tumours: revised RECIST guideline (version 1.1). Eur J Cancer 2009;45(2):228–47 doi 10.1016/j.ejca.2008.10.026.

20. Jarraya H, Mirabel X, Taieb S, Dewas S, Tresch E, Bonodeau F, et al. Image-based response assessment of liver metastases following stereotactic body radiotherapy with respiratory tracking. Radiat Oncol 2013;8:24 doi 10.1186/1748-717X-8-24.

21. Bruix J, Sherman M, Llovet JM, Beaugrand M, Lencioni R, Burroughs AK, et al. Clinical Management of Hepatocellular Carcinoma. Conclusions of the Barcelona-2000 EASL Conference. Journal of Hepatology 2001;35(3):421–30 doi 10.1016/S0168-8278(01)00130-1.

22. Gillmore R, Stuart S, Kirkwood A, Hameeduddin A, Woodward N, Burroughs AK, et al. EASL and mRECIST responses are independent prognostic factors for survival in hepatocellular cancer patients treated with transarterial embolization. Journal of Hepatology 2011;55(6):1309–16 doi 10.1016/j.jhep.2011.03.007.

23. Kim BK, Kim KA, Park JY, Ahn SH, Chon CY, Han K-H, et al. Prospective comparison of prognostic values of modified Response Evaluation Criteria in Solid Tumours with European Association for the Study of the Liver criteria in hepatocellular carcinoma following chemoembolisation. European Journal of Cancer 2013;49(4):826–34 doi 10.1016/j.ejca.2012.08.022.

24. Tacher V, Lin M, Duran R, Yarmohammadi H, Lee H, Chapiro J, et al. Comparison of Existing Response Criteria in Patients with Hepatocellular Carcinoma Treated with Transarterial Chemoembolization Using a 3D Quantitative Approach. Radiology 2016;278(1):275–84 doi 10.1148/radiol.2015142951.

25. Ghosn M, Derbel H, Kharrat R, Oubaya N, Mulé S, Chalaye J, et al. Prediction of overall survival in patients with hepatocellular carcinoma treated with Y-90 radioembolization by imaging response criteria. Diagn Interv Imaging 2021;102(1):35–44 doi 10.1016/j.diii.2020.09.004.

26. Benson AB, D’Angelica MI, Abbott DE, Anaya DA, Anders R, Are C, et al. Hepatobiliary cancers, version 2.2021, NCCN clinical practice guidelines in oncology. Journal of the National Comprehensive Cancer Network 2021;19(5):541–65.

27. Enderling H, Alfonso JCL, Moros E, Caudell JJ, Harrison LB. Integrating Mathematical Modeling into the Roadmap for Personalized Adaptive Radiation Therapy. Trends Cancer 2019;5(8):467–74 doi 10.1016/j.trecan.2019.06.006.

28. Koay EJ, Odisio BC, Javle M, Vauthey J-N, Crane CH. Management of unresectable intrahepatic cholangiocarcinoma: how do we decide among the various liver-directed treatments? Hepatobiliary Surgery and Nutrition 2017;6(2):105–16.

29. Dogra P, Ramírez JR, Butner JD, Peláez MJ, Chung C, Hooda-Nehra A, et al. Translational Modeling Identifies Synergy between Nanoparticle-Delivered miRNA-22 and Standard-of-Care Drugs in Triple-Negative Breast Cancer. Pharmaceutical Research 2022 doi 10.1007/s11095-022-03176-3.

30. Wang CX, Elganainy D, Zaid MM, Butner JD, Agrawal A, Nizzero S, et al. Mass transport model of radiation response: calibration and application to chemoradiation for pancreatic cancer. International Journal of Radiation Oncology*Biology*Physics 2022 doi 10.1016/j.ijrobp.2022.04.044.

31. Galeaz C, Totis C, Bisio A. Radiation Resistance: A Matter of Transcription Factors. Front Oncol 2021;11:662840 doi 10.3389/fonc.2021.662840.

32. Schwarz K, Dobiasch S, Nguyen L, Schilling D, Combs SE. Modification of radiosensitivity by Curcumin in human pancreatic cancer cell lines. Scientific Reports 2020;10(1):3815 doi 10.1038/s41598-020-60765-1.

33. Li Q, Hu Y, Xi M, He L, Zhao L, Liu M. Sorafenib modulates the radio sensitivity of hepatocellular carcinoma cells in vitro in a schedule-dependent manner. BMC Cancer 2012;12(1):485 doi 10.1186/1471-2407-12-485.

34. Zaid M, Elganainy D, Dogra P, Dai A, Widmann L, Fernandes P, et al. Imaging-Based Subtypes of Pancreatic Ductal Adenocarcinoma Exhibit Differential Growth and Metabolic Patterns in the Pre-Diagnostic Period: Implications for Early Detection. 2020;10(2629) doi 10.3389/fonc.2020.596931.

35. He LN, Zhang X, Li H, Chen T, Chen C, Zhou Y, et al. Pre-Treatment Tumor Growth Rate Predicts Clinical Outcomes of Patients With Advanced Non-Small Cell Lung Cancer Undergoing Anti-PD-1/PD-L1 Therapy. Front Oncol 2020;10:621329 doi 10.3389/fonc.2020.621329.

36. Ten Berge D, Hurkmans DP, den Besten I, Kloover JS, Mathijssen RHJ, Debets R, et al. Tumour growth rate as a tool for response evaluation during PD-1 treatment for non-small cell lung cancer: a retrospective analysis. ERJ Open Res 2019;5(4) doi 10.1183/23120541.00179-2019.

37. Osorio B, Yegya-Raman N, Kim S, Simone CB, 2nd, Theodorou Ross C, Deek MP, et al. Clinical significance of pretreatment tumor growth rate for locally advanced non-small cell lung cancer. Ann Transl Med 2019;7(5):95 doi 10.21037/atm.2019.02.14.

38. Dromain C, Pavel ME, Ruszniewski P, Langley A, Massien C, Baudin E, et al. Tumor growth rate as a metric of progression, response, and prognosis in pancreatic and intestinal neuroendocrine tumors. BMC Cancer 2019;19(1):66 doi 10.1186/s12885-018-5257-x.

39. Lamarca A, Crona J, Ronot M, Opalinska M, Lopez Lopez C, Pezzutti D, et al. Value of Tumor Growth Rate (TGR) as an Early Biomarker Predictor of Patients’ Outcome in Neuroendocrine Tumors (NET)-The GREPONET Study. Oncologist 2019;24(11):e1082–e90 doi 10.1634/theoncologist.2018-0672.

40. Chapiro J, Wood LD, Lin M, Duran R, Cornish T, Lesage D, et al. Radiologic-pathologic analysis of contrast-enhanced and diffusion-weighted MR imaging in patients with HCC after TACE: diagnostic accuracy of 3D quantitative image analysis. Radiology 2014;273(3):746–58 doi 10.1148/radiol.14140033.

41. Tacher V, Lin M, Chao M, Gjesteby L, Bhagat N, Mahammedi A, et al. Semiautomatic volumetric tumor segmentation for hepatocellular carcinoma: comparison between C-arm cone beam computed tomography and MRI. Acad Radiol 2013;20(4):446–52 doi 10.1016/j.acra.2012.11.009.

42. Tacher V, Lin M, Duran R, Yarmohammadi H, Lee H, Chapiro J, et al. Comparison of Existing Response Criteria in Patients with Hepatocellular Carcinoma Treated with Transarterial Chemoembolization Using a 3D Quantitative Approach. Radiology 2016;278(1):275–84 doi 10.1148/radiol.2015142951.

43. Zaid M, Chaudhury B, Varadhachary GR, Katz MHG, Herman JM, Tamm EP, et al. Discovery and validation of a quantitative, stromal-associated imaging biomarker of pancreatic ductal adenocarcinoma (PDAC). Journal of Clinical Oncology 2018;36(4_suppl):228- doi 10.1200/JCO.2018.36.4_suppl.228.

44. Chapiro J, Duran R, Lin M, Schernthaner R, Lesage D, Wang Z, et al. Early survival prediction after intra-arterial therapies: a 3D quantitative MRI assessment of tumour response after TACE or radioembolization of colorectal cancer metastases to the liver. European Radiology 2015;25(7):1993–2003 doi 10.1007/s00330-015-3595-5.

45. Chapiro J, Lin M, Duran R, Schernthaner RE, Geschwind JF. Assessing tumor response after loco-regional liver cancer therapies: the role of 3D MRI. Expert review of anticancer therapy 2015;15(2):199–205 doi 10.1586/14737140.2015.978861.

46. Namjan A, Techasen A, Loilome W, Sa-Ngaimwibool P, Jusakul A. ARID1A alterations and their clinical significance in cholangiocarcinoma. PeerJ 2020;8:e10464 doi 10.7717/peerj.10464.

47. Zhao S, Xu Y, Wu W, Wang P, Wang Y, Jiang H, et al. ARID1A Variations in Cholangiocarcinoma: Clinical Significances and Molecular Mechanisms. Front Oncol 2021;11:693295 doi 10.3389/fonc.2021.693295.

48. Park PC, Choi GW, M. Zaid M, Elganainy D, Smani DA, Tomich J, et al. Enhancement pattern mapping technique for improving contrast-to-noise ratios and detectability of hepatobiliary tumors on multiphase computed tomography. Medical Physics 2020;47(1):64–74 doi 10.1002/mp.13769.

49. Cazoulat G, Elganainy D, Anderson BM, Zaid M, Park PC, Koay EJ, et al. Vasculature-Driven Biomechanical Deformable Image Registration of Longitudinal Liver Cholangiocarcinoma Computed Tomographic Scans. Advances in Radiation Oncology 2020;5(2):269–78 doi 10.1016/j.adro.2019.10.002.

50. Sen A, Anderson BM, Cazoulat G, McCulloch MM, Elganainy D, McDonald BA, et al. Accuracy of deformable image registration techniques for alignment of longitudinal cholangiocarcinoma CT images. Medical Physics 2020;47(4):1670–9 doi 10.1002/mp.14029.

