## Supplementary Information for "Measurable imaging-based changes in enhancement of intrahepatic cholangiocarcinoma after radiotherapy reflect physical mechanisms of response"

**Table S1.** List of mathematical model constants.

| **Parameter** | $\boldsymbol{\delta}_{\mathbf{max}}$ **(d^-1^)** | $\boldsymbol{D}_{\boldsymbol{50}}$ **(Gy)** | $\boldsymbol{k}$ **(d^-1^)** |
| --- | --- | --- | --- |
| **Definition** | Radiation-induced *in vitro* tumor death rate | Half-maximal inhibitory radiation dose | Decay rate constant of radiation |
| **Value** | 0.05 | 1 | 0.1 |
| **Reference** | (33) | (32) | Fixed |

**Table S2.** Results of univariate and multivariable Cox analyses examining associations with overall survival and local control

| **Attribute** | | **Overall survival** | | | | **Local control** | | | |
| --- | --- | --- | --- | --- | --- | --- | --- | --- | --- |
|  |  | Univariate analysis | | Multivariable analysis | | Univariate analysis | | Multivariable analysis | |
|  |  | *HR [95% CI]* | *P-value* | *HR [95% CI]* | *P-value* | *HR [95% CI]* | *P-value* | *HR [95% CI]* | *P-value* |
| **Age*** | | 1.01 [0.99-1.02] | 0.412 | 1.01 [0.99-1.03] | 0.370 | 1.00 [0.99-1.02] | 0.846 | 0.99 [0.97-1.01] | 0.265 |
| **Stage** | I-III | Reference | | Reference | | Reference | | Reference | |
|  | IV | 1.54 [1.07-2.22] | 0.021* | 1.58 [1.08-2.33] | 0.019* | 1.49 [1.05-2.13] | 0.027* | 1.33 [0.77-2.29] | 0.302 |
| **Radiation BED (Gy)** | | 0.99 [0.98-1.00] | 0.038* | 0.99 [0.98-1.00] | 0.106 | 0.97 [0.96-0.99] | <0.001* | 0.97 [0.96-0.99] | <0.001* |
| **Change in viable volume after RT** | | 1.06 [1.02-1.09] | <0.001* | 1.05 [1.02-1.08] | 0.001* | 1.04 [1.00-1.08] | 0.083 | 1.04 [0.99-1.08] | 0.088 |

* Significant at 5% level; BED, biologically effective dose; HR, hazard ratio; RT, radiotherapy

**Table S3**. Mutational correlates of changes in quantitative viability volume.

|  |  | Univariate associations with percent change in viability volume | | |
| --- | --- | --- | --- | --- |
| **Mutation** | **Frequency (%)** | **Coefficient** | **95% confidence interval** | ***P*-value** |
| *TP53* | 23 (24%) | -0.46 | -2.94 to +2.02 | 0.714 |
| *IDH1* | 22 (23%) | -1.11 | -3.62 to +1.40 | 0.381 |
| *BAP1* | 18 (19%) | -0.66 | -3.36 to +2.05 | 0.630 |
| *ARID1A* | 17 (18%) | +2.94 | +0.24 to +5.64 | 0.033* |
| *IDH2* | 16 (17%) | -1.97 | -4.78 to +0.83 | 0.166 |
| *PIK3CA* | 11 (12%) | -0.64 | -3.95 to +2.68 | 0.704 |
| *MLL* | 11 (12%) | -1.93 | -5.22 to +1.36 | 0.248 |
| None | 10 (11%) | -0.74 | -4.20 to +2.71 | 0.670 |
| *CDKN2A* | 10 (11%) | -1.01 | -4.46 to +2.44 | 0.563 |
| *FGFR2* | 10 (11%) | -0.82 | -4.27 to +2.63 | 0.638 |
| *BRCA2* | 6 (6%) | -1.39 | -5.74 to +2.96 | 0.527 |

*Significant at 5% level

**Table S4.** Results of model-derived univariate and multivariable Cox analyses examining associations with overall survival and local control

| **Attribute** | | **Overall survival** | | | | **Local control** | | | |
| --- | --- | --- | --- | --- | --- | --- | --- | --- | --- |
|  |  | Univariate analysis | | Multivariable analysis | | Univariate analysis | | Multivariable analysis | |
|  |  | *HR [95% CI]* | *P-value* | *HR [95% CI]* | *P-value* | *HR [95% CI]* | *P-value* | *HR [95% CI]* | *P-value* |
| **Age** | | 1.01 [0.99-1.02] | 0.412 | 1.01 [0.99-1.03] | 0.217 | 1.00 [0.99-1.02] | 0.846 | 0.99 [0.97-1.01] | 0.463 |
| **Stage** | I-III | Reference | | Reference | | Reference | | Reference | |
|  | IV | 1.54 [1.07-2.22] | 0.021* | 1.76 [1.20-2.58] | 0.004* | 1.49 [1.05-2.13] | 0.027* | 1.59 [0.91-2.80] | 0.104 |
| **Radiation BED (Gy)** | | 0.99 [0.98-1.00] | 0.038* | 0.99 [0.99-1.00] | 0.085 | 0.97 [0.96-0.99] | <0.001* | 0.98 [0.97-0.99] | 0.001* |
| **Responder to RT, as measured by binary γ classifier** | | 0.40 [0.26-0.61] | <0.001* | 0.38 [0.25-0.59] | <0.001* | 0.29 [0.15-0.56] | <0.001* | 0.31 [0.15-0.61] | 0.001* |

**Table S5**. Patient-specific parameter estimates from model fitting.

| **ID** | **BVF** | **Dose (Gy)** | **Fractions** | $\boldsymbol{\gamma}$ **(d^-1^)** | $\boldsymbol{\alpha}$ | $\boldsymbol{D}_{\boldsymbol{50}}^{\mathbf{eff}}$ **(Gy)** |
| --- | --- | --- | --- | --- | --- | --- |
| *Non-responders* | | | | | | |
| 13 | 0.07799432 | 45.00 | 18 | 0.012421956 | 0.99659163 | 12.7104453 |
| 31 | 0.07247838 | 66.00 | 20 | 0.023477941 | 0.99737797 | 13.7025993 |
| 112 | 0.04615563 | 60.00 | 10 | 0.01778064 | 0.96234669 | 19.2965193 |
| 21 | 0.00177045 | 60.00 | 30 | 0.019559198 | 0.99420827 | 544.47449 |
| 137 | 0.00579637 | 67.50 | 15 | 0.008091366 | 0.92959505 | 120.049104 |
| 128 | 0.01918761 | 67.50 | 15 | 0.011158743 | 0.9982137 | 51.7502094 |
| 30 | 0.04589509 | 50.40 | 28 | 0.003334406 | 0.7997814 | 11.7569455 |
| 20 | 0.08944069 | 67.50 | 15 | 0.026166673 | 0.95925048 | 10.1330549 |
| 47 | 0.00270992 | 67.50 | 15 | 0.016318099 | 0.62959695 | 41.324015 |
| 50 | 0.01104323 | 67.50 | 15 | 0.03923977 | 0.95904256 | 75.2929347 |
| 9 | 0.03432314 | 50.00 | 25 | 0.021685969 | 0.55237105 | 6.44019958 |
| 27 | 0.00388853 | 50.40 | 28 | 0.038830315 | 0.96190727 | 208.162745 |
| 87 | 0.0833425 | 67.50 | 15 | 0.023448865 | 0.60328482 | 2.42251573 |
| 79 | 0.23069746 | 62.50 | 25 | 0.024809211 | 0.99926756 | 11.9768626 |
| 94 | 0.00264899 | 60.00 | 15 | 0.017304386 | 0.86232591 | 6.49349607 |
| 60 | 0.11423671 | 100.00 | 25 | 0.008042537 | 0.96011096 | 297.939597 |
| 43 | 0.11719623 | 60.00 | 25 | 0.004194441 | 0.7581092 | 5.0800166 |
| 139 | 0.00836511 | 60.00 | 10 | 0.03901676 | 0.96251165 | 99.918323 |
| 42 | 0.10806822 | 63.00 | 28 | 0.014748589 | 0.98413351 | 8.9324411 |
| 90 | 0.06370257 | 90.00 | 15 | 0.007012511 | 0.25637659 | 2.02575146 |
| 12 | 0.00519061 | 35.00 | 14 | 0.018936857 | 0.48451001 | 12.7937966 |
| 22 | 0.04838186 | 50.40 | 28 | 0.026235171 | 0.98998668 | 20.0514951 |
| 135 | 0.0835033 | 60.00 | 15 | 0.019473305 | 0.97180116 | 2680.36515 |
| 40 | 0.00029671 | 58.05 | 15 | 0.023005784 | 0.97742988 | 11.322937 |
| 129 | 0.06119078 | 60.00 | 15 | 0.0204226 | 0.77990168 | 8.83620213 |
| 104 | 0.11085992 | 45.00 | 15 | 0.033850517 | 0.94273339 | 7.9528522 |
| 103 | 0.07334992 | 60.00 | 15 | 0.019337022 | 0.84066185 | 8.99114948 |
| 49 | 0.0024485 | 75.00 | 25 | 0.0128136 | 0.71712796 | 74.5576872 |
| 105 | 0.34751781 | 60.00 | 15 | 0.005751742 | 0.44999711 | 1.60901093 |
| 68 | 0.00648026 | 62.50 | 25 | 0.019388187 | 0.5373104 | 14.9918089 |
| 92 | 0.00724094 | 100.00 | 25 | 0.012465777 | 0.68715422 | 29.5563565 |
| 4 | 0.16528926 | 35.00 | 14 | 0.019761548 | 0.92116258 | 5.24957076 |
| 69 | 0.02570231 | 60.00 | 15 | 0.00910353 | 0.93943899 | 31.1698118 |
| 2 | 0.04020513 | 60.00 | 15 | 0.018723179 | 0.98411446 | 23.6345212 |
| 46 | 0.01334217 | 50.40 | 28 | 0.016501777 | 0.98325022 | 69.722251 |
| 6 | 0.01725963 | 50.40 | 28 | 0.017061084 | 0.86482263 | 33.4699587 |
| 55 | 0.00113015 | 50.40 | 28 | 0.011916929 | 0.97137721 | 728.648794 |
| 144 | 0.01396914 | 67.50 | 15 | 0.002664674 | 0.17660069 | 2.12600505 |
| 145 | 0.17921495 | 67.50 | 15 | 0.017899214 | 0.95320701 | 20.9435979 |
| 17 | 0.04112427 | 50.00 | 10 | 0.022892848 | 0.92706627 | 4.92233956 |
| 41 | 0.01591906 | 75.00 | 25 | 0.024619205 | 0.91640614 | 44.4399887 |
| 154 | 0.99641455 | 60.00 | 15 | 0.029452025 | 0.99276476 | 1.00357227 |
| 141 | 0.12874787 | 67.50 | 15 | 0.012715741 | 0.97700181 | 7.40944332 |
| 16 | 0.00838283 | 50.40 | 28 | 0.020191798 | 0.98895242 | 113.153486 |
| 134 | 0.07287524 | 67.50 | 15 | 0.009016426 | 0.50018611 | 3.7061389 |
| 81 | 0.00159669 | 60.00 | 15 | 0.010842641 | 0.48560986 | 22.8109519 |
| 88 | 0.66145251 | 75.00 | 25 | 0.009678461 | 0.99954964 | 1.51154294 |
| 52 | 0.01068877 | 67.50 | 15 | 0.000884951 | 0.16204668 | 2.08643923 |
| 109 | 0.006108 | 67.50 | 15 | 0.015357557 | 0.66104654 | 29.0819786 |
| 118 | 0.16003337 | 75.00 | 15 | 0.014315002 | 0.48604141 | 2.43661343 |
| 149 | 0.002429 | 67.50 | 15 | 0.011308411 | 0.89309316 | 232.241781 |
| 76 | 0.00224309 | 60.00 | 25 | 0.02444551 | 0.97362189 | 351.24053 |
| 24 | 0.00979671 | 35.00 | 14 | 0.035366148 | 0.95634161 | 83.4091001 |
| 100 | 0.01264141 | 67.50 | 15 | 0.009535871 | 0.9634229 | 67.4176544 |
| 33 | 0.00072882 | 66.00 | 20 | 0.005877602 | 0.33151534 | 10.9670298 |
| 150 | 0.2987784 | 67.50 | 15 | 0.01264119 | 0.97281304 | 156.806505 |
| 19 | 0.0055371 | 50.00 | 25 | 0.009277875 | 0.26528113 | 1.37778025 |
| 133 | 0.02461277 | 58.05 | 15 | 0.027890415 | 0.93877285 | 32.3843207 |
| 153 | 0.03259827 | 67.50 | 15 | 0.025686552 | 0.98980573 | 29.6243346 |
| 82 | 0.17916428 | 75.00 | 25 | 0.004816327 | 0.36742259 | 1.88092853 |
| 146 | 0.04713366 | 67.50 | 15 | 0.00936016 | 0.88434689 | 14.9016891 |
| 63 | 0.10953596 | 60.00 | 15 | 0.021776966 | 0.89240487 | 7.19621699 |
| 65 | 0.13577839 | 50.40 | 28 | 0.020098278 | 0.98160287 | 7.09930652 |
| 122 | 0.00520296 | 67.50 | 15 | 0.000647157 | 0.61114243 | 24.8711847 |
| 3 | 0.05452136 | 55.80 | 31 | 0.023960433 | 0.67731019 | 7.1735847 |
| 152 | 0.00617781 | 60.00 | 15 | 0.026593166 | 0.97405101 | 141.853347 |
| 132 | 0.02519507 | 60.00 | 15 | 0.018452542 | 0.97563592 | 36.2855646 |
| 48 | 0.01666667 | 67.50 | 15 | 0.009644173 | 0.76640212 | 23.0557306 |
| 36 | 0.01035844 | 58.05 | 15 | 0.004633459 | 0.67821825 | 22.1852184 |
| 45 | 0.21135417 | 58.05 | 15 | 0.004163898 | 0.58367299 | 2.47726777 |
| 136 | 0.00688092 | 67.50 | 15 | 0.022695576 | 0.83542366 | 64.0444518 |
| 113 | 0.00872242 | 67.50 | 15 | 0.027364988 | 0.99122284 | 109.973376 |
| 25 | 0.72924623 | 66.00 | 20 | 0.003223236 | 0.36656787 | 1.12270567 |
| 89 | 0.5119814 | 69.00 | 23 | 0.023637694 | 0.99825751 | 1.9509188 |
| 51 | 0.11104272 | 100.00 | 25 | 0.005271851 | 1 | 9.00554301 |
| 61 | 0.09519451 | 100.00 | 25 | 0.012403182 | 0.74665509 | 5.78928086 |
| 119 | 0.01053763 | 75.00 | 25 | 0.018745561 | 0.85650535 | 49.3777266 |
| 15 | 0.02242656 | 50.40 | 28 | 0.012636994 | 0.62246463 | 10.6314351 |
| 10 | 0.04335679 | 50.40 | 28 | 0.004806409 | 0.98973217 | 22.3330683 |
| 108 | 0.05571062 | 60.00 | 15 | 0.023236151 | 0.87388259 | 12.4710488 |
| 56 | 0.01236345 | 58.05 | 15 | 0.002611393 | 0.15497412 | 1.97546337 |
| 151 | 0.00371411 | 60.00 | 15 | 0.038656344 | 0.98485938 | 247.372312 |
| 53 | 0.0045646 | 58.05 | 15 | 0.003321598 | 0.46173979 | 12.0433401 |
| 91 | 0.39708495 | 62.50 | 25 | 0.006101767 | 0.3217911 | 1.34609509 |
| 142 | 0.00361902 | 67.50 | 15 | 0.027305595 | 0.98517012 | 254.216455 |
| 114 | 0.01609272 | 60.00 | 15 | 0.013212812 | 0.95668641 | 51.962897 |
| 84 | 0.12777976 | 90.00 | 15 | 0.014629688 | 0.97969082 | 7.50569467 |
| 148 | 0.07654146 | 70.00 | 10 | 0.018621062 | 0.99258059 | 12.8180637 |
| 99 | 0.81706435 | 67.50 | 15 | 0.003899251 | 0.97793438 | 1.21844973 |
| 66 | 0.29851656 | 75.00 | 15 | 0.004712987 | 0.75071396 | 2.47826945 |
| 73 | 0.03605785 | 60.00 | 15 | 2.22E-14 | 2.22E-14 | 1 |
| 131 | 0.23601996 | 67.50 | 15 | 0.034977585 | 0.98440328 | 4.14258409 |
| 93 | 0.54428518 | 67.50 | 15 | 0.013342566 | 0.82489931 | 1.65164433 |
| 111 | 0.1825054 | 50.40 | 28 | 3.20E-07 | 5.30E-05 | 1.00009008 |
| 26 | 0.44744432 | 60.00 | 30 | 0.006623545 | 0.82988558 | 1.9491563 |
| 143 | 0.29197314 | 67.50 | 15 | 0.008177467 | 0.98735307 | 3.37206022 |
| 117 | 0.12689467 | 67.50 | 15 | 0.018516179 | 0.6699144 | 3.98672426 |
| 18 | 0.26844637 | 54.00 | 30 | 0.016997717 | 0.98635544 | 3.658891 |
| 96 | 0.16312087 | 100.00 | 25 | 0.00662783 | 0.83642379 | 4.55696578 |
| 101 | 0.16251477 | 40.00 | 8 | 0.019609162 | 0.82916983 | 4.51132227 |
| 72 | 0.26508213 | 100.00 | 25 | 0.001101611 | 0.25669833 | 1.40610346 |
| 130 | 0.0124251 | 67.50 | 15 | 0.004476005 | 0.70587586 | 22.1405164 |
| 70 | 0.08013852 | 58.05 | 15 | 0.000980271 | 0.6706622 | 5.43440753 |
| 5 | 0.13767425 | 53.82 | 26 | 0.005816391 | 0.54607016 | 2.95288704 |
| 11 | 0.21929388 | 50.40 | 28 | 0.007303363 | 0.44139971 | 1.95375773 |
| 34 | 0.10026906 | 58.05 | 15 | 0.005768652 | 0.99913452 | 9.95333447 |
| 38 | 0.02465608 | 58.05 | 15 | 0.00332297 | 0.96599215 | 35.7592739 |
| 147 | 0.43499542 | 67.50 | 15 | 2.78E-11 | 3.29E-09 | 1 |
| 58 | 0.12383097 | 60.00 | 3 | 0.005221937 | 0.33690457 | 2.02129625 |
| 59 | 0.13910741 | 58.05 | 15 | 0.004323968 | 0.81279574 | 4.96914035 |
| 116 | 0.05769978 | 67.50 | 15 | 0.001070663 | 0.17548703 | 1.649673 |
| 7 | 0.01847469 | 50.00 | 20 | 0.010652378 | 0.99966825 | 54.0564867 |
| 97 | 0.38370975 | 67.50 | 15 | 0.016339756 | 0.95492753 | 2.4960147 |
| 206 | 0.01136824 | 50.00 | 10 | 0.005175845 | 0.53085476 | 10.7682326 |
| 71 | 0.14220428 | 100.00 | 25 | 0.00718315 | 0.96196203 | 6.5292884 |
| 138 | 0.24470301 | 67.50 | 15 | 0.015770966 | 0.92881649 | 3.69693558 |
| 23 | 0.01700528 | 50.40 | 28 | 1.92E-08 | 0.72993851 | 19.5688411 |
| 98 | 0.51191195 | 60.00 | 15 | 0.00477564 | 0.95187108 | 1.89150988 |
| 86 | 0.44037851 | 100.00 | 25 | 0.011344384 | 0.60508927 | 1.64254395 |
| 125 | 0.07459766 | 67.50 | 15 | 5.17E-04 | 0.08571948 | 1.24919262 |
| *Responders* | | | | | | |
| 83 | 0.04547727 | 75.00 | 15 | 0.016391745 | 0.96723988 | 19.8716897 |
| 95 | 0.0411449 | 75.00 | 25 | 1.12E-08 | 1.21E-06 | 1.00000386 |
| 35 | 0.44002976 | 58.05 | 15 | 0.001091456 | 0.26182242 | 1.23977933 |
| 78 | 0.00871853 | 75.00 | 15 | 0.017358661 | 0.96520876 | 97.2528528 |
| 64 | 0.28862788 | 54.00 | 15 | 0.005046195 | 0.75344441 | 2.55038124 |
| 77 | 0.16662958 | 58.05 | 15 | 0.008695386 | 0.99593732 | 5.95780278 |
| 140 | 0.24239564 | 60.00 | 15 | 0.00115026 | 0.79115817 | 3.06858549 |
| 62 | 0.32406848 | 62.50 | 25 | 0.006005388 | 0.92615235 | 2.83938893 |
| 121 | 0.20660714 | 67.50 | 15 | 0.006498083 | 0.98339524 | 4.71501221 |
| 126 | 0.04641257 | 60.00 | 15 | 0.00064515 | 0.19660181 | 1.82869742 |
| 115 | 0.01179651 | 67.50 | 15 | 0.002917065 | 0.36515377 | 5.05948663 |
| 57 | 0.04953209 | 67.50 | 15 | 0.00443175 | 0.571105 | 5.56361097 |
| 102 | 0.06511118 | 75.00 | 15 | 0.00100971 | 0.44939396 | 3.41299784 |
| 127 | 0.03386542 | 75.00 | 15 | 0.003861217 | 0.97040987 | 26.7139945 |
| 85 | 0.05013092 | 67.50 | 15 | 0.01050832 | 0.48321146 | 4.2474066 |
| 110 | 0.06140933 | 67.50 | 15 | 0.000573679 | 0.3518812 | 2.66930107 |
| 75 | 0.21083055 | 90.00 | 15 | 2.22E-14 | 2.38E-14 | 1 |
| 39 | 0.04932646 | 58.05 | 15 | 0.00080625 | 0.23654001 | 2.03769363 |
| 124 | 0.43195231 | 67.50 | 15 | 0.01277676 | 0.96578588 | 2.24952569 |
| 120 | 0.0054023 | 75.00 | 15 | 0.002345251 | 0.37022616 | 6.90969244 |
| 74 | 0.32161616 | 50.00 | 4 | 1.18E-09 | 2.28E-07 | 1.00000026 |
| 29 | 0.10619077 | 60.00 | 30 | 0.003956203 | 0.90100087 | 7.54217632 |
| 123 | 0.01851815 | 100.00 | 25 | 0.003402841 | 0.63131855 | 12.4078846 |
| 32 | 0.01546715 | 62.50 | 25 | 1.59E-08 | 8.02E-06 | 1.00003345 |
| 8 | 0.04634772 | 60.00 | 20 | 0.005949564 | 0.467473 | 4.2033514 |
| 37 | 0.04118214 | 58.05 | 15 | 0.001860962 | 0.21037411 | 1.95627268 |
| 80 | 0.59900896 | 58.05 | 15 | 0.012975548 | 0.81744448 | 1.52032314 |
| 14 | 0.02046649 | 50.40 | 28 | 0.008344734 | 0.91377048 | 34.9396692 |
| 44 | 0.0091203 | 58.05 | 15 | 6.38E-12 | 0.81895071 | 46.8438035 |
| 67 | 0.12971349 | 67.50 | 15 | 2.96E-12 | 3.13E-10 | 1 |
| 1 | 0.06660777 | 75.00 | 15 | 0.014769171 | 0.37291064 | 2.74612358 |
| 107 | 0.03285714 | 50.00 | 4 | 0.001931126 | 0.54400324 | 6.41147861 |
| 28 | 0.1248472 | 66.00 | 20 | 2.22E-14 | 2.34E-12 | 1 |
| 54 | 0.11673307 | 67.50 | 15 | 0.001133144 | 0.43944044 | 2.56987928 |

**Table S6**. Simulation results for % change in enhancement with variable start times of radiotherapy relative to initial diagnosis.

| Dose (Gy) | Fractions | % Change in enhancement 60 days after RT termination for responders | % Change in enhancement 60 days after RT termination for non-responders | % Change in enhancement 60 days after RT termination for responders (treatment starting 7 days sooner) | % Change in enhancement 60 days after RT termination for non-responders (treatment starting 7 days sooner) |
| --- | --- | --- | --- | --- | --- |
| 66 | 17 | -80.1934 | -14.1647 | -80.8396 | -21.1907 |
| 50.4 | 28 | -81.68304751 | -1.172930667 | -82.5492961 | -12.39104821 |
| 100 | 25 | -87.78252413 | -34.94887724 | -88.13633226 | -44.95534017 |
| 67.5 | 15 | -78.89447161 | -6.549318715 | -79.7954345 | -19.66668669 |
| 70 | 10 | -75.43303272 | -8.646556621 | -77.58700738 | -16.75765757 |
| 50 | 4 | -65.96343546 | 7.388706353 | -69.52638886 | 1.901452617 |

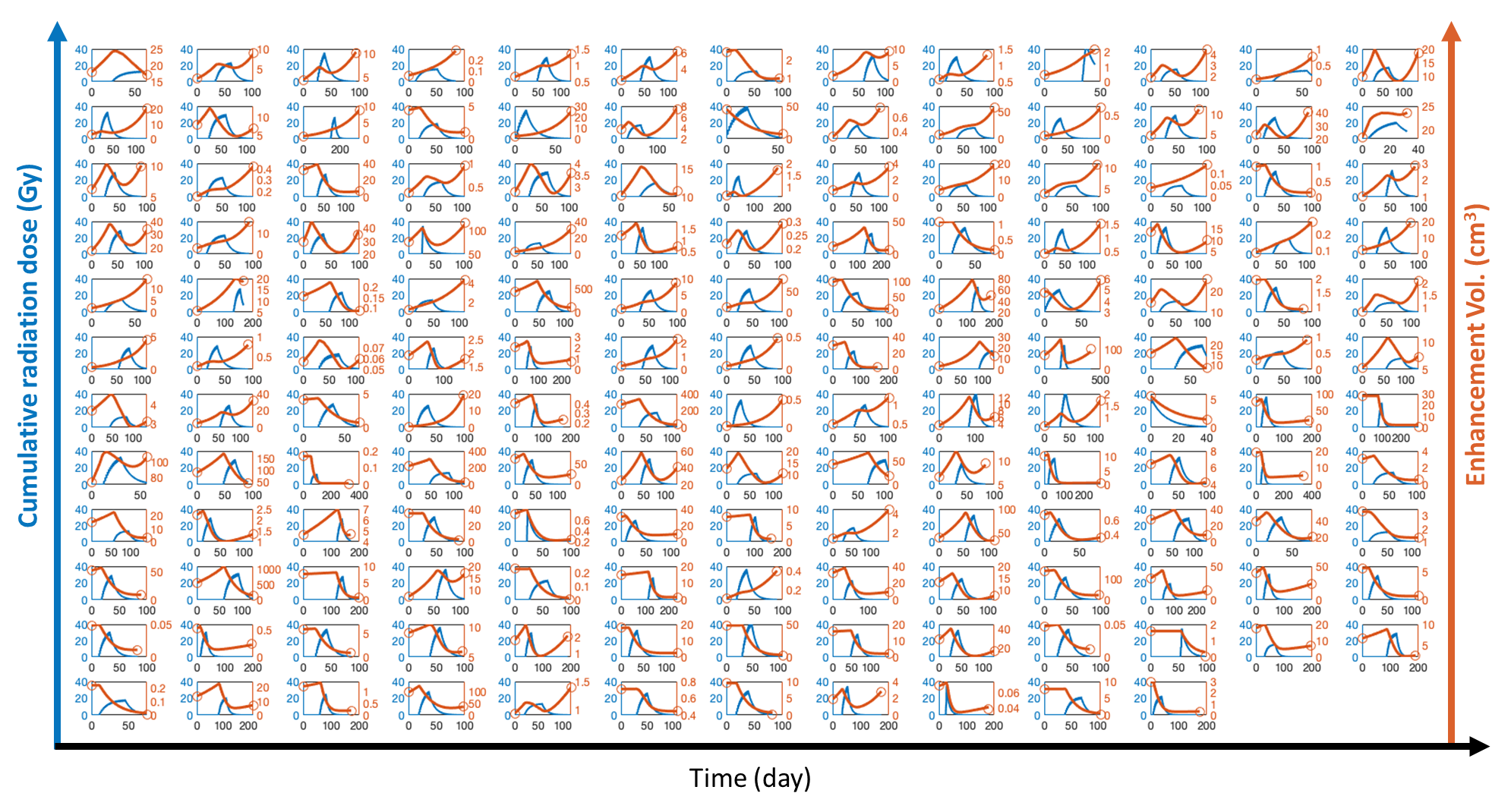

**Figure S1**. Patient-specific model fits.

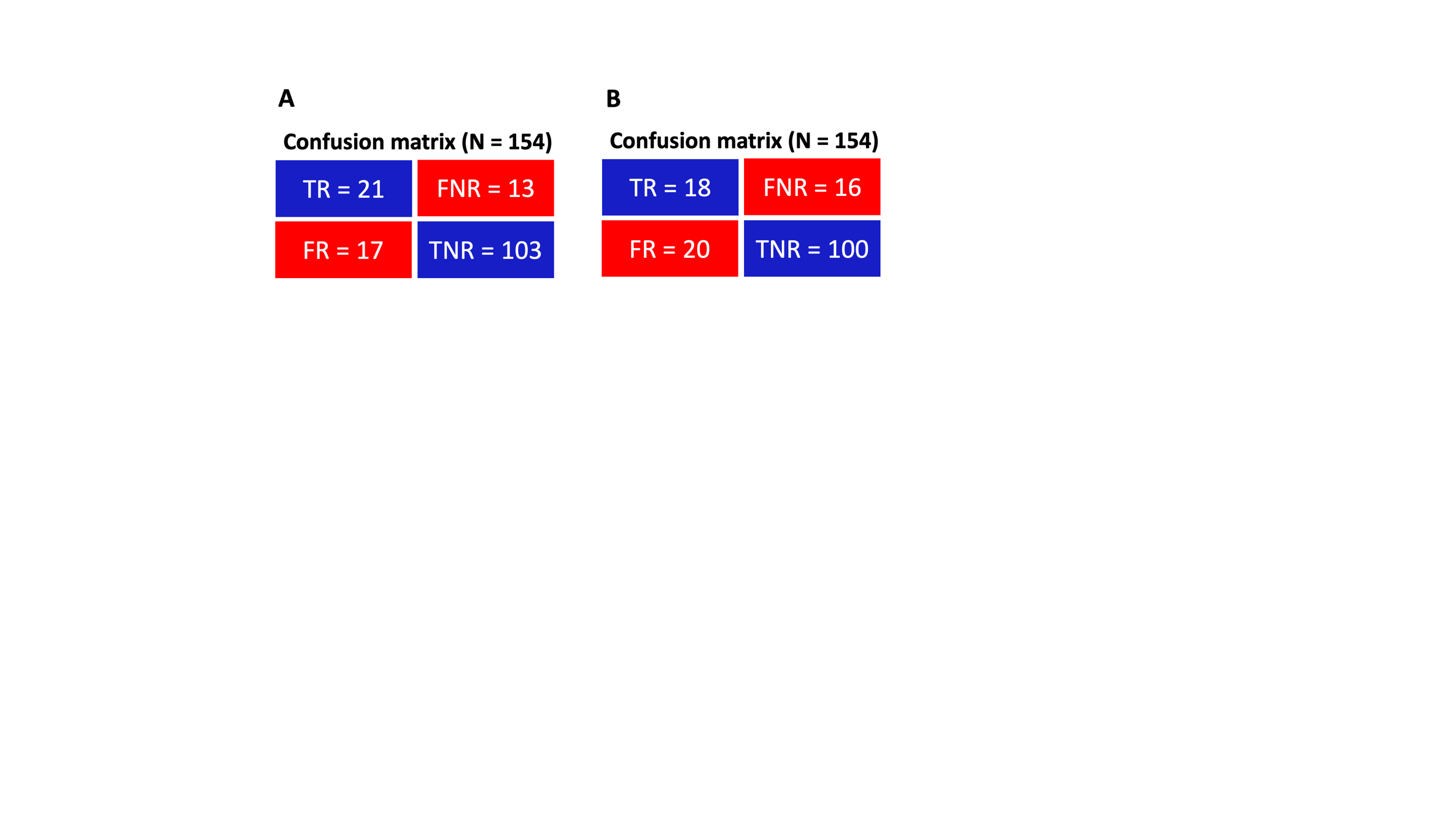

**Figure S2.** Confusion matrices corresponding to binary classification and LOOCV shown in A) Figure 4B and B) Figure 4C, respectively.
